# Global dynamics of antibiotic resistance: a modeling study of the spread of endemic and emerging resistance in *E. coli*

**DOI:** 10.1101/2025.09.25.25335519

**Authors:** Eve Rahbe, Eugenio Valdano, David RM Smith, Didier Guillemot, Simon Cauchemez, Philippe Glaser, Lulla Opatowski

## Abstract

Antibiotic resistance (ABR) is a global threat, yet its dynamics strongly vary across countries. Drivers of such heterogeneity remain unclear, limiting efforts to mitigate emergence and worldwide spread of multi-resistant clones. We developed a mechanistic modeling framework to analyze extended-spectrum β-lactamase-producing *Escherichia coli* (ESBL-EC) dynamics globally accounting for antibiotic consumption and international mobility. Calibrated with data from 39 countries (2006-2019), the model is used to explore the global dissemination of potential emerging multi-resistant clones, like carbapenem-resistant *E. coli* (CR-EC). The model estimated heterogeneous within-country transmission rates and an excess risk of ABR acquisition under antibiotics of 9.83 (95%CrI:9.78-9.89). Simulations of potential CR-EC dissemination showed that mobility patterns drive early resistance dynamics, while long-term dynamics are mostly predicted by within-country transmission and antibiotic use. Our findings suggest that preventing introduction at the country-level is key to mitigate emerging threats but reducing within-country transmission and selection pressure is fundamental to avoid fixation.

## Introduction

In 2021, an estimated 1.14 million deaths worldwide were attributed to antibiotic-resistant bacteria, and antibiotic-resistant *Escherichia coli* are among the leading causes of death.^1^ *E. coli* is both a commensal symbiont of the intestinal microbiome and an opportunistic pathogen causing severe intestinal and extra-intestinal infections.^2^ Globally, there is great concern regarding an increasing prevalence of *E. coli* producing extended-spectrum β-lactamase (ESBL), which confer resistance to most β-lactam antibiotics, including penicillins, third generation cephalosporins (3GC) and monobactams.^3^ The World Health Organization identified 3GC-resistant Enterobacterales, including ESBL-producing *E. coli* (ESBL-EC), as critical priority pathogens for the development of novel therapeutic strategies.^4^

ESBL production was first observed in a clinical isolate of *Klebsiella pneumoniae* in Europe in 1983.^5^ Following their rapid expansion, first in hospital-acquired infections and later in community-acquired infections,^3^ ESBL-EC are now globally endemic^6^ but with large geographic variation: reported 3GC prevalence among *E. coli* bloodstream infections varies from 5.3% in Norway to 90% in Ivory Coast,^7^ while asymptomatic ESBL-EC colonization prevalence varies from an estimated 6.0% in Europe to 27% in South-East Asia.^8^ This global expansion of ESBL-producing Enterobacterales has led to increased use of carbapenems, a last resort β-lactam,^9^ and, in turn, to a rise of carbapenem resistance in *K. pneumoniae* in some regions.^10^ There is now concern that the acquisition of carbapenem resistance among rapidly spreading strains of *E. coli* could lead to their further global dissemination, mimicking the epidemiology of ESBL-EC.^11,12^

Little is known about factors driving heterogeneity in ESBL-EC prevalence across countries.^13^ At an individual level, a known risk factor for ESBL-EC infection and colonization is antibiotic exposure.^14,15^ Antibiotic use is highly country-specific, with global consumption data suggesting stark heterogeneity in levels and trends across countries,^16^ although a spatiotemporal analysis suggests that country-level antibiotic sales are only associated with the evolution of resistance in some drug-bug pairs and noticeably not in ESBL-EC.^6^ Another important individual risk factor for ESBL-EC colonization is travel to highly endemic areas, such as South-East Asia or northern Africa,^17^ and international mobility is one of the world’s fastest growing economic sectors.^18^ In order to implement effective control measures for endemic bacteria like ESBL-EC and prevent the further dissemination of emerging multidrug-resistant clones, there is a clear need to better understand which factors have driven the rapid yet heterogeneous global spread of ESBL-EC clones.

Despite progress in implementing and standardizing robust surveillance programs,^19^ global antibiotic resistance data are still patchy. While hospital-based infection surveillance is routine in some regions, with some facilities even conducting routine asymptomatic colonization swabs, many other regions have only limited or biased sampling.^20^ Moreover, colonization surveillance in the community relies on point prevalence studies with limited spatiotemporal coverage.^8^ In this context, mathematical models allows to analyze available epidemiological data and evaluate the likelihood of various proposed causal mechanisms driving observed ESBL-EC dynamics. Through simulations, such models can further help to explore potential scenarios for the hypothetical spread of emerging resistances in *E. coli*.^21^

In this study, we propose a multi-country analysis of available data on ESBL-EC at the global level, using a mathematical model of transmission, colonization and infection. The model calibrated on historical data of resistance trends in bloodstream infections is then used to assess hypothetical scenarios for the international spread of emerging antibiotic-resistant bacteria, such as a putative transmissible carbapenem-resistant *E. coli* (CR-EC) clone.

## Material & Methods

### 1. Model description

We developed a deterministic compartmental model that formalizes the transmission of antibiotic-resistant *E. coli* in large, homogeneously mixed human populations at the country level. The model accounts for mechanisms specific to *E. coli*’s within-host ecology and epidemiology. Using a metapopulation approach, multiple countries are structured as interconnected subpopulations linked via a mobility network that allows for bacterial transmission and acquisition through travel (Figure 1, supplementary Equation 1).

**Figure 1.**
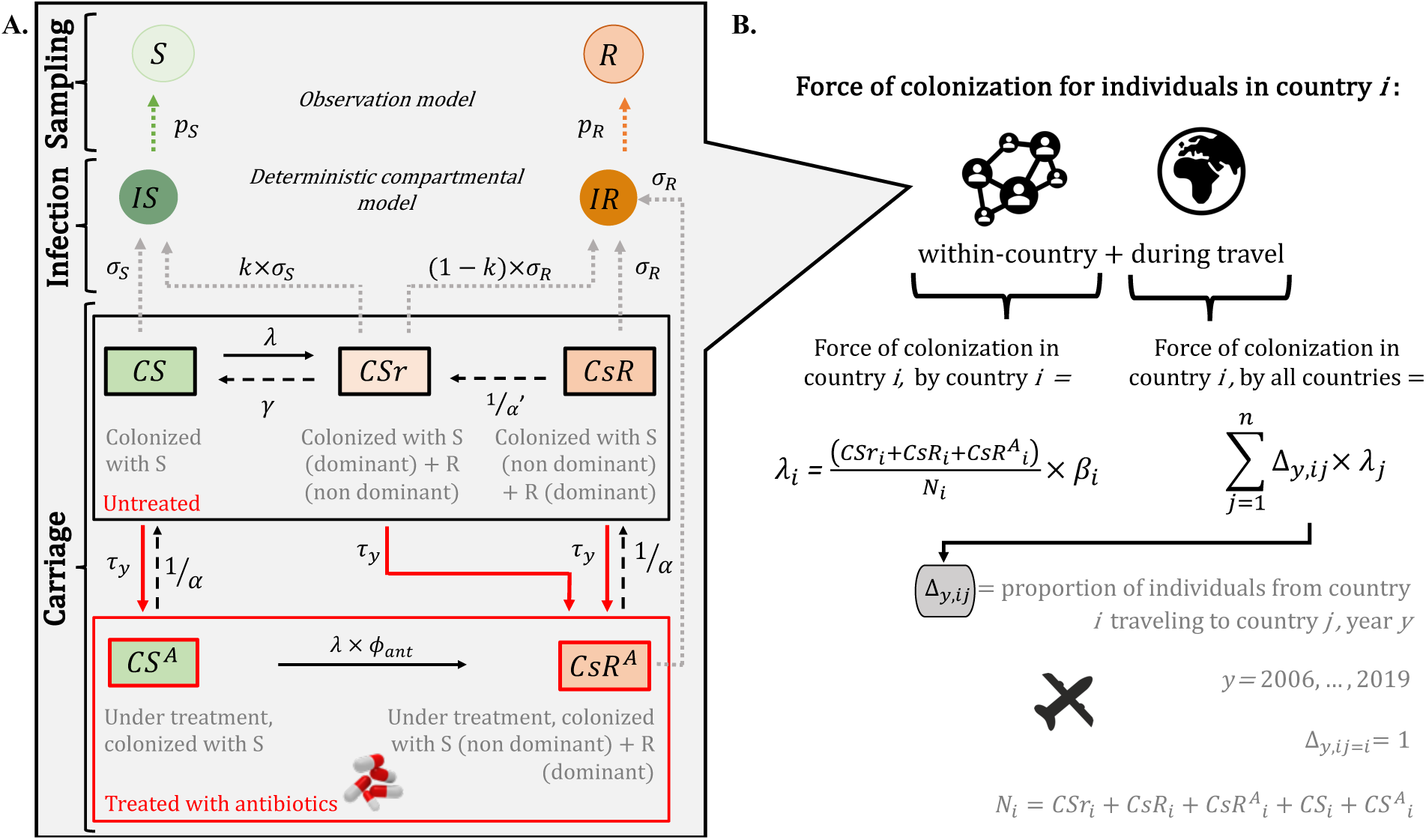
Antibiotic-resistant *E. coli* transmission model. **(A)** Schematic of the deterministic compartmental model and observation model for within-country transmission of antibiotic-resistant *E. coli*. **Compartments**: ***CS*** individuals colonized with antibiotic-Sensitive strain; ***CSr*** individuals colonized with antibiotic-Sensitive strain and sub-dominant antibiotic-Resistant strain; ***CsR*** individuals colonized with antibiotic-Sensitive strain and dominant antibiotic-Resistant strain; ***CS^A^*** individuals colonized with antibiotic-Sensitive strain and exposed to antibiotics; and ***CsR^A^***individuals colonized with antibiotic-Sensitive strain and dominant antibiotic-Resistant strain and exposed to antibiotics; ***IS*** infected individuals with antibiotic-sensitive bacteria; ***IR*** infected individuals with antibiotic-resistant bacteria; ***S*** observed infected individuals with antibiotic-sensitive bacteria within monitoring system; ***R*** observed infected individuals with antibiotic-resistant bacteria within monitoring system. **Parameters**: Force of colonization ***λ***; natural clearance rate **γ**; duration of antibiotic treatment ***α***; duration of microbiome dysbiosis ***α***′; individual antibiotic exposure rate ***τ*** at year *y*; excess risk of acquisition of antibiotic-resistant strain under antibiotic treatment ***ϕ_ant_***; colonization-to-infection progression rate for antibiotic-sensitive and antibiotic-resistant strains ***σ_S_*** and ***σ_R_***; proportion of co-colonized individuals developing infection by antibiotic-sensitive strain ***k***; proportions of infections with antibiotic-sensitive and antibiotic-resistant strain reported in the monitoring system data compared to national estimates ***p_S_*** and ***p_R_***. **(B)** Meta-population structure and force of colonization including between-country transmission through travel.

#### Colonization by *E. coli*

The model considers three distinct states of *E. coli* colonization among individuals recently exposed or not to antibiotics. As *E. coli* is a commensal intestinal bacteria present in over 90% of the human population,^22^ we assume that in the absence of antibiotic treatment everyone in the population is colonized by antibiotic-sensitive *E. coli*, and that some individuals are further co-colonized by transmissible antibiotic-resistant *E. coli*. We also assume that strains colonize the gut either dominantly or sub-dominantly. Thus, individuals can be colonized solely by an antibiotic-sensitive strain (CS); colonized dominantly by an antibiotic-sensitive strain, and sub-dominantly by an antibiotic-resistant strain (CSr); and colonized dominantly by a resistant strain, and sub-dominantly by a sensitive strain (CsR) (Figure 1, A). The sub-dominant antibiotic-resistant strain is naturally cleared after an average of 1/γ days. Thus, non-exposed individuals solely acquire sub-dominant resistant strains and can only become colonized by dominant resistant strains upon antibiotic exposure. Antibiotic exposure is assumed to last an average of α days and to induce microbiome dysbiosis, which increases the risk of exposed individuals acquiring dominantly antibiotic-resistant strains by a coefficient ϕ_ant_. Dysbiosis is assumed to last for an average period of α’ days after the end of antibiotic exposure.^23^ In addition, as antibiotics are assumed to directly promote dominant colonization by antibiotic-resistant bacteria (described by CsR^A^), subdominant antibiotic-resistant colonization is not considered among antibiotic-exposed individuals.

#### Infection by *E. coli*

Most strains responsible for extra-intestinal *E. coli* infections primarily colonize the human gut.^2^ In the model, colonized individuals develop extra-intestinal infections at rate σ_S_ if colonized dominantly by antibiotic-sensitive *E. coli* and rate σ_R_ if colonized dominantly by antibiotic-resistant *E. coli*. If an individual is co-colonized by a dominant sensitive strain and a sub-dominant resistant strain, we assume that the dominant strain has a higher chance of being the cause of infection, such that infection by the antibiotic-sensitive strain occurs at rate k×σ_S_ and by the resistant strain at rate (1-k)×σ_R_ with k assumed to be higher than 0.5 (Figure 1, A). Finally, as antibiotics are effective against sensitive strains, antibiotic-exposed individuals colonized only by a sensitive strain (CS^A^) are assumed not to develop *E. coli* infection.

#### Within-country transmission of antibiotic-resistant *E. coli*

We assume that acquisition of antibiotic-resistant *E. coli* in country *i* occurs through interactions with colonized individuals (dominant or sub-dominant) at a transmission rate β_i_. This reflects direct or indirect inter-human transmission routes. The force of colonization *λ_i_*(*t*) represents the colonization risk exerted on CS individuals and depends on the proportion of CSr, CsR and CsR^A^ individuals (Figure 1, B). Independent of colonization status, in year *y* individuals are exposed to antibiotic treatment at an average rate of τ_i,y_.

#### Between-country transmission of antibiotic-resistant *E. coli*

We consider that the *n* countries included in the analysis are connected through a mobility network matrix of size *n*×*n*. Matrix values are defined as the proportion of travelers from country *i* travelling to country *j* each day of year *y*, Δ*_y_*_,*i*j_(Figure 1, A). The mobility matrix is parameterized using data from the Knowledge Center for Migration and Demography,^24^ which reports the estimated cumulative annual number of travelers between country pairs for 2011-2016 (supplementary Figure 1). Individuals can travel to several countries during the same year. For simplicity, we consider exportation of resistance as negligible. Importing risk only come from travels of CS individuals who may further be colonized by an antibiotic-resistant strain in the visited country by contacts with CSr, CsR and CsR^A^ individuals. Travelers from country *i* to country *j* are assumed to mix homogeneously with the destination country population, acquiring or transmitting according to the local force of colonization.

#### Stochastic observation model

A stochastic observation model is added on top of the transmission model to relate model outcomes (the number of antibiotic-sensitive and -resistant infections) to observed infection data from a monitoring system (Figure 1, A). See details in supplementary Equation 2.

### 2. Data analysis and model comparison

#### Data

We calibrated the model to observed global trends of ESBL-EC colonization and bloodstream infections obtained from various data sources.

##### Number of antibiotic-sensitive and -resistant *E. coli* infections reported over 2006-2019

Annual time series over 2006-2019 of the reported number of bloodstream infections by *E. coli* sensitive or resistant to 3GC in *n* = 39 countries were provided by the Antimicrobial Testing Leadership and Surveillance (ATLAS) reporting system, and used to calibrate the proportions of infections by sensitive versus resistant bacteria over time. ATLAS is a hospital monitoring program led by Pfizer which collects antibiotic susceptibilities of clinical isolates from different bacterial species, offering antibiotic resistance rates trends over a wide range of countries.^25^ Here, we restricted our analyses to bloodstream infections, for which hospitalizations are generally systematic. This choice was made to minimize surveillance bias towards infections by antibiotic-resistant bacteria as less serious infections due to antibiotic-sensitive bacterial infections may be treated in the community and reports in hospital databases may depend on the country healthcare organization. Resistance to 3GC was used as a proxy for ESBL production in *E. coli*.

##### Proportion of asymptomatic colonization by antibiotic-resistant *E. coli* over 2006-2019

To the best of our knowledge, the meta-analysis by Bezabih and colleagues provides the most recent and comprehensive estimates of fecal ESBL-EC colonization in healthy volunteers.^8^ Here we use averaged proportion values over the period 2003-2018 by country, depicted in supplementary Figure 2.

##### Incidence of *E. coli* bloodstream infections observed over 2006-2019

To our knowledge, there is no temporal estimate of the incidence of *E. coli* bloodstream infections worldwide. We review the literature to extract annual incidences of *E. coli* bloodstream infections in blood for different regions of the world and for one year as detailed in supplementary Table 3.

#### Statistical inference framework

##### Model parametrization

Parameter values were either, fixed using values from the literature or provided by available surveillance or monitoring databases; or estimated by Bayesian inference from empirical data (parameters in supplementary Table 1). As the model represents the selection of ESBL-EC by antibiotics, we inform the antibiotic treatment rate using yearly national consumption of *β*-lactams, including broad-spectrum penicillins, monobactams and cephalosporins from the IQVIA MIDAS database (references in supplementary Table 1). Demographic data from the World Bank were used for all countries included in the analysis (supplementary Table 2).

##### Bayesian inference

We used a Bayesian inference approach to estimate the posterior distributions of unknown parameters and calibrate the model to available data described above for the different countries. The expression for likelihood *L* is presented in supplementary Equation 3. The Markov Chain Monte Carlo (MCMC) method was used, implemented via an Adaptative Metropolis algorithm in the *modMCMC* function (FME package, R software).^26^ Visual inspection was done to assess convergence (supplementary Figure 7 and 8 for the selected model).

##### Model comparison

Different models, with increasing complexity and country specificity, were independently fitted to the global data described above. Models were then compared with the aim of identifying which mechanisms best explain global ESBL-EC colonization and infection dynamics. For each model, a distinct combination of parameters was estimated simultaneously. The six models explored are presented in Table 1, and the model comparison procedure (using Deviance Information Criteria) and selection are further described in supplementary section 4. For the selected model, we computed mean absolute errors (MAE) between observed and expected values of antibiotic resistance proportions at each annual timepoint to assess model fit.

**Table 1.** Description of the models evaluated. The mechanisms included are described for the six evaluated models. Each model has a distinct number of parameters estimated, with *n* = the number of countries included in the analysis. The orange boxes indicate the new mechanism introduced in the evaluated model compared with the previous one. *Travel*: The cross (x) indicates models for which travel is accounted for, i.e. that the mobility matrix represents the estimated proportion of travelers per year and per country (absence of cross indicates that travel is not accounted for, i.e. that the travel matrix is a null matrix with a diagonal of 1). *Effect of antibiotic on R acquisition*: by default, the parameter ***ϕ_ant_*** is set to 1. If ***ϕ_ant_*** is estimated, it is a country-independent value. *Other parameters*: estimates were either country-independent (***a***, ***β*** and ***σ_S_***) or country-dependent (***a_i_***, ***β_i_***and ***σ_S i_***). R: antibiotic-resistant bacteria. S: antibiotic-sensitive bacteria.

| Mechanisms | Travel | Estimated parameters |  |  |  | Number of estimated parameters | Number of data points from time series |
| --- | --- | --- | --- | --- | --- | --- | --- |
|  |  | Effect of antibiotic on R acquisition | Colonized-to-infected progression rate ratio for R vs S | Transmission rate of R | Colonized-to-infected progression rate for S |  |  |
| **Model 1** | - | $\phi_{ant} = 1$ | $a$ | $\beta$ | $\sigma_S$ | 3 | 14 years × $n$ countries |
| **Model 2** | x | $\phi_{ant} = 1$ | $a$ | $\beta$ | $\sigma_S$ | 3 | |
| **Model 3** | x | $\phi_{ant}$ | $a$ | $\beta$ | $\sigma_S$ | 4 | |
| **Model 4** | x | $\phi_{ant}$ | $a_i$ | $\beta$ | $\sigma_S$ | $1 \times n + 3$ | |
| **Model 5** | x | $\phi_{ant}$ | $a_i$ | $\beta_i$ | $\sigma_S$ | $2 \times n + 2$ | |
| **Model 6** | x | $\phi_{ant}$ | $a_i$ | $\beta_i$ | $\sigma_{S i}$ | $3 \times n + 1$ | |

#### Sensitivity analyses

Multivariable sensitivity analyses were performed to quantify the impact of uncertainty around the input parameters on the model’s outcomes using Latin hypercube sampling. Partial rank correlation coefficients were then used to quantify and rank the impact of each parameter (supplementary Table 4, Figures 4 and 5).

### 3. Simulation of emergence scenarios

To assess the risk of a hypothetical emerging CR-EC clone to spread globally, a simulation study was carried out using the multi-country deterministic compartmental model previously calibrated on ESBL-EC. The model parameters used are those of the selected model, corresponding to median values derived from posterior distributions.

#### Initialization

Emergence is modeled by introducing 20 individuals colonized by CR-EC into the CSr compartment at model initialization (t = 0 days) in one index country. This is conceptualized as representing an initial emergence event in a hospital with subsequent spread into the community. We run independent simulations initializing each country as the index country and assessed global transmission dynamics over 20 years. A “control” case without importation through travel was also assessed for each country, representing within-country dynamics in absence of travel.

#### Antibiotic consumption

Here, carbapenem exposure is assumed to exert selection pressure on the emerging CR-EC clone. Data on carbapenem consumption per country-year were taken from the MIDAS IQVIA database.^27^ As data were available for 14 years (2006-2019) out of the 20 years of simulation, we performed linear extrapolation for missing years (supplementary Figure 3).

#### Emerging clone scenarios

Different scenarios based on plausible biological hypotheses were tested to explore a range of characteristics of hypothetical CR-EC clones, described in Table 2.

**Table 2.** Scenarios evaluated for the global spread of hypothetical CR-EC clones.

|  | Name | Description |
| --- | --- | --- |
| Scenario s1 | “Base-case CR-EC” | This scenario represents the worldwide spread of a hypothetical *E. coli* clone that has acquired carbapenem resistance. Model's transmission parameters are assumed to be the same as those for ESBL-EC, such as $\beta_i(s1) = \beta_i$. |
| Scenario s2 | “Fitness cost” | In this scenario, we assume that the CR-EC clone has a lower transmission rate than the clone in scenario 1, due to a fitness cost associated with the acquisition of this new resistance. Model's transmission parameters are modified, with a 10% reduction in transmission such that $\beta_i(s2) = \beta_i \times 0.9$. |
| Scenario s3 | “Super-bug” | In this scenario, we assume that the emerging CR-EC clone has a higher transmission rate than the clone in scenario 1. This could represent an already circulating super-transmissible *E. coli* clone that independently acquires carbapenem resistance. Model's transmission parameters are modified, with a 10% increase in transmission such that $\beta_i(s3) = \beta_i \times 1.1$. |
| Scenario s4 | “Multi-drug resistance” | In this scenario, we assume that the emerging Cr-EC clone is multi-drug resistant. Model's transmission parameters are assumed to be the same as those for ESBL-EC such as $\beta_i(s4) = \beta_i$. However, we consider that other antibiotic classes may co-select the clone. Parameter values $\tau_{y,i}$ are therefore provided here by both per capita treatments per day of carbapenems and β-lactams (broad-spectrum penicillins, monobactams and cephalosporins). |

#### Outcomes

The primary outcome is predicted CR-EC infection incidence.

## Results

### Trends in ESBL-EC infections and antibiotic use

ESBL-EC rates and trends reported by ATLAS showed heterogeneity across the 39 countries included in the analysis for both infections and colonization. From 2006-2019, average rates of 3GC resistance among bloodstream isolates ranged from 4.2% (7.6% standard deviation, sd) in Austria to 83% (12.8% sd) in Pakistan (Figure 2).

**Figure 2.**
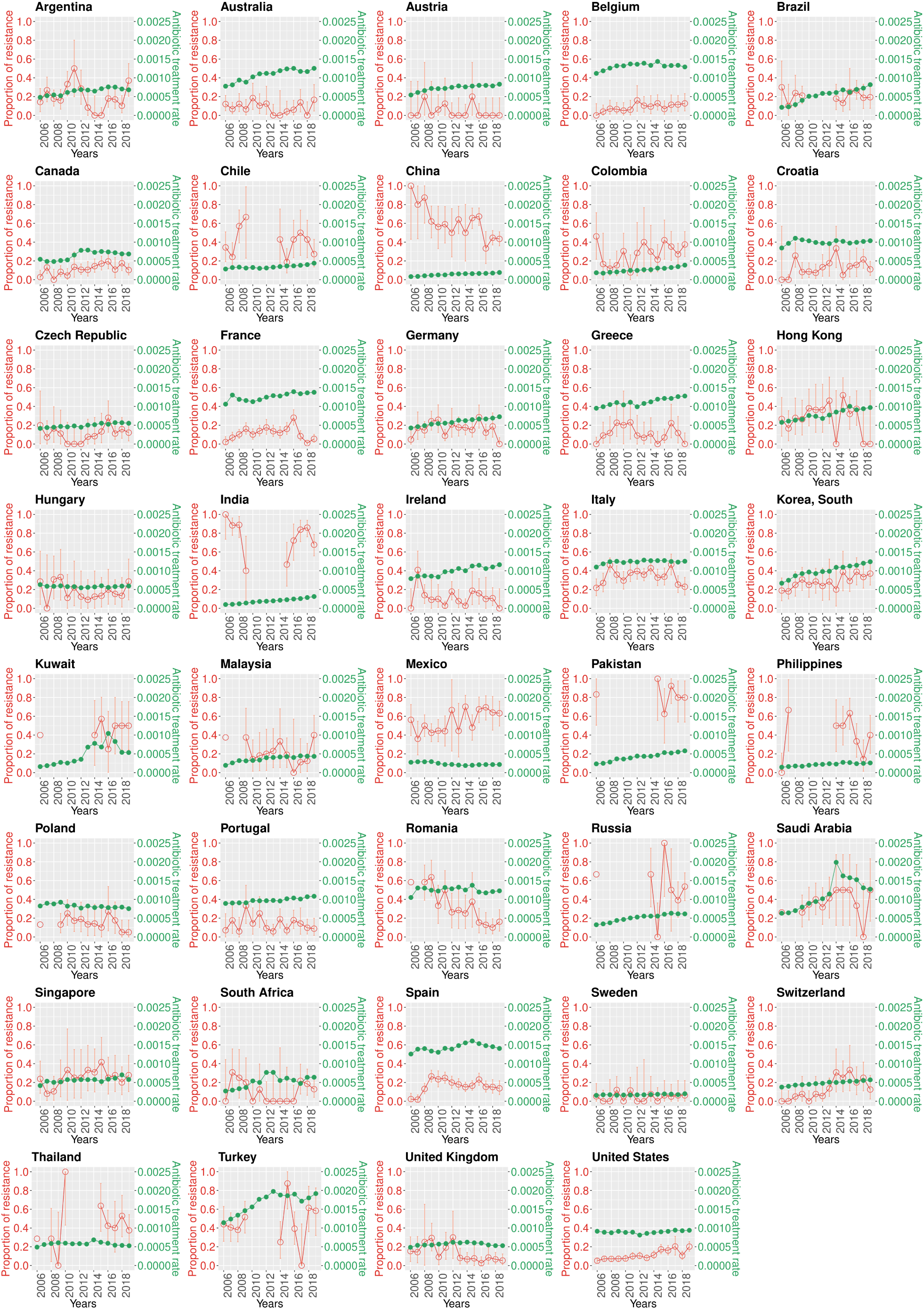
Trends of ESBL-producing *E. coli* and β-lactam antibiotic consumption in 39 countries over 2006-2019. Annual trends by country for ESBL-producing *E. coli* proportion in bloodstream infection isolates from the ATLAS database in red and *β*-lactam antibiotic consumption in treatment courses per individual per day from the IQVIA MIDAS database in green. Here resistance to third generation cephalosporins is used as a proxy for ESBL production. 95% confidence intervals around antibiotic resistance proportions are calculated using Bayesian probability interval estimates with Jeffrey priority. β-lactam antibiotics include broad-spectrum penicillins, monobactams and cephalosporins.

Large heterogeneities were also observed in national β-lactam use reported by IQVIA MIDAS. In 2006, rates ranged from 8.3×10^−5^ individual treatments per day in China to 126 ×10^−5^ in Spain. Over 2006-2019, daily treatment rates were stable or increasing everywhere, except for Mexico, where rates decreased slightly (Figure 2). Carbapenem consumption was estimated to be either stable or increasing in 38 out of 39 countries from 2006-2019 (supplementary Figure 3).

### Model selection and calibration

The different models listed in Table 1 were independently fitted to the above-mentioned spatio-temporal datapoints. After model comparison, model 6 was selected (DIC = 4991). Corresponding posterior distributions for estimated parameters showed high between-country heterogeneity (Figure 3, B). The median ESBL-EC transmission rate (*β_i_*) had an approximate twofold range, from 7.05×10^−3^ (7.03×10^−3^; 7.08×10^−3^ 95% credible interval) day^−1^ in Spain to 1.621×10^−2^ (1.616×10^−2^; 1.627×10^−2^) day^−1^ in Thailand. Individuals exposed to antibiotics had an estimated 9.83 (9.78; 9.89) times higher risk of acquiring ESBL-EC than non-exposed individuals (*ϕ_ant_*). The probability of progressing from colonization to infection for sensitive strains (σ_Si_) ranged from 1.01×10^−6^ (1×10^−6^; 1.04×10^−6^) day^−1^ for India to 1.68×10^−6^ (1.4×10^−6^; 1.9×10^−6^) day^−1^ for the United Kingdom. The probability ratio of progressing from colonization to infection for resistant compared to sensitive strains (*a_i_*) ranged from 2.5 (2.42; 2.58) in Austria to 90.74 (90.67; 90.8) in Pakistan. Pakistan, India (68.11), Russia (67.36) and Hungary (40.86) were identified as outliers (*a_i_* > 30). Estimated β_i_ median values were positively correlated with mean national temperatures (*r* = 0.64, p-value <1×10^−5^) and negatively associated with mean national gross domestic product (*r* = −0.36, p-value = 0.02) (supplementary Figure 10).

**Figure 3.**
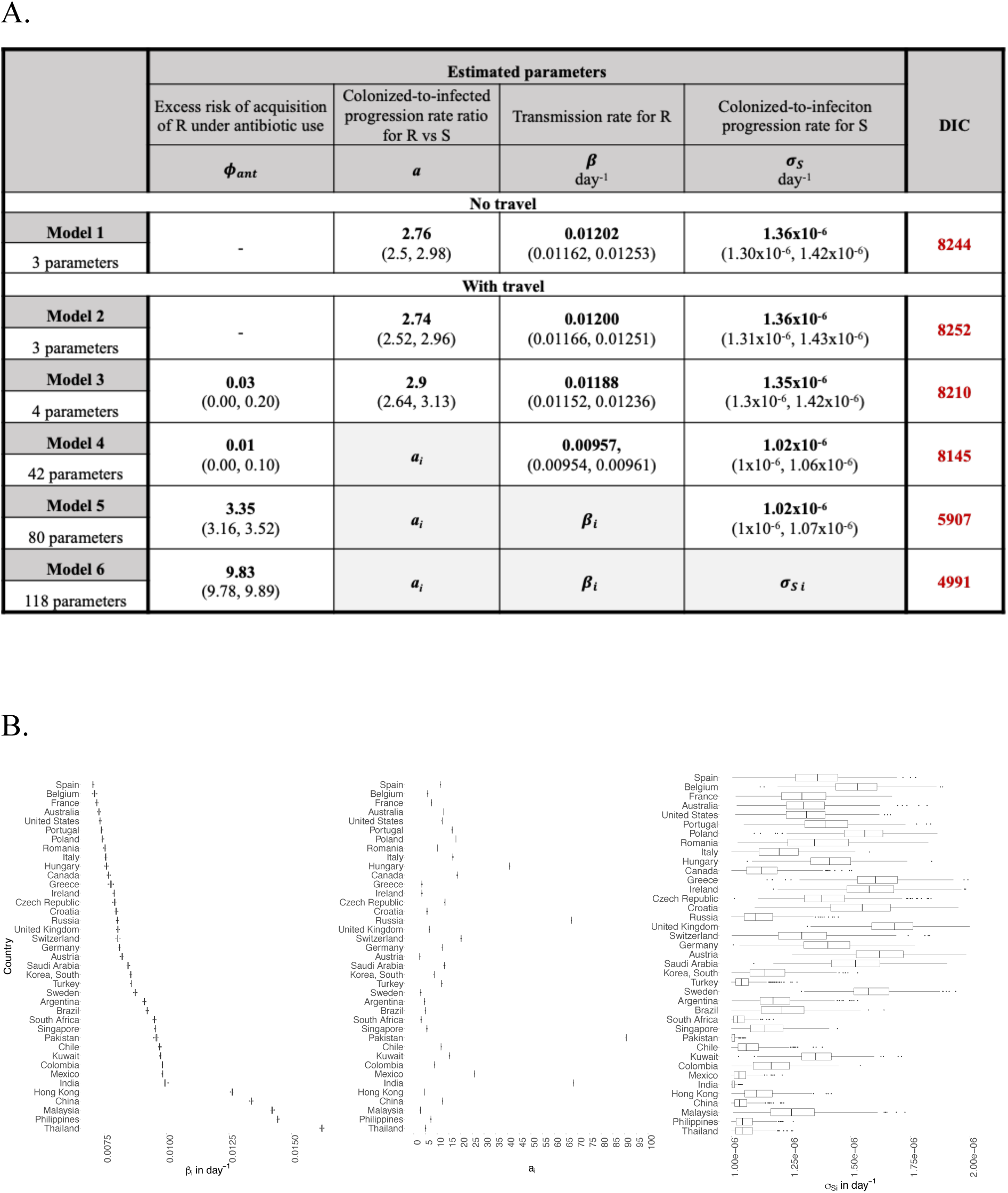
Model comparison and parameter estimates for the selected model. **(A)** Comparison of the six models tested, with associated median and 95% credibility intervals from posterior distributions of parameters and deviance information criteria (DIC), with lowest DIC privileged. **(B)** Posterior distributions for country-dependent parameters *β_i_*, *a_i_* and *σ_S i_*for the selected model (model 6). Countries are ordered by increasing median *β_i_* values.

The selected model overall reproduced the observed country-level dynamics of ESBL-EC rates among bloodstream isolates (Figure 4). The MAE ranged from 2% in Belgium to 27% in Russia and it was lower than 10% in 24 over the 39 analyzed countries. The model did not reproduce decreasing temporal dynamics observed in China and Romania (MAE = 16% and 17%, respectively). Wide 95% prediction intervals around the model’s median trajectories were observed in Kuwait and Russia, driven by small sample sizes in the ATLAS data (n = 55 and 78 for Kuwait and Russia, respectively, over 2006-2019).

**Figure 4.**
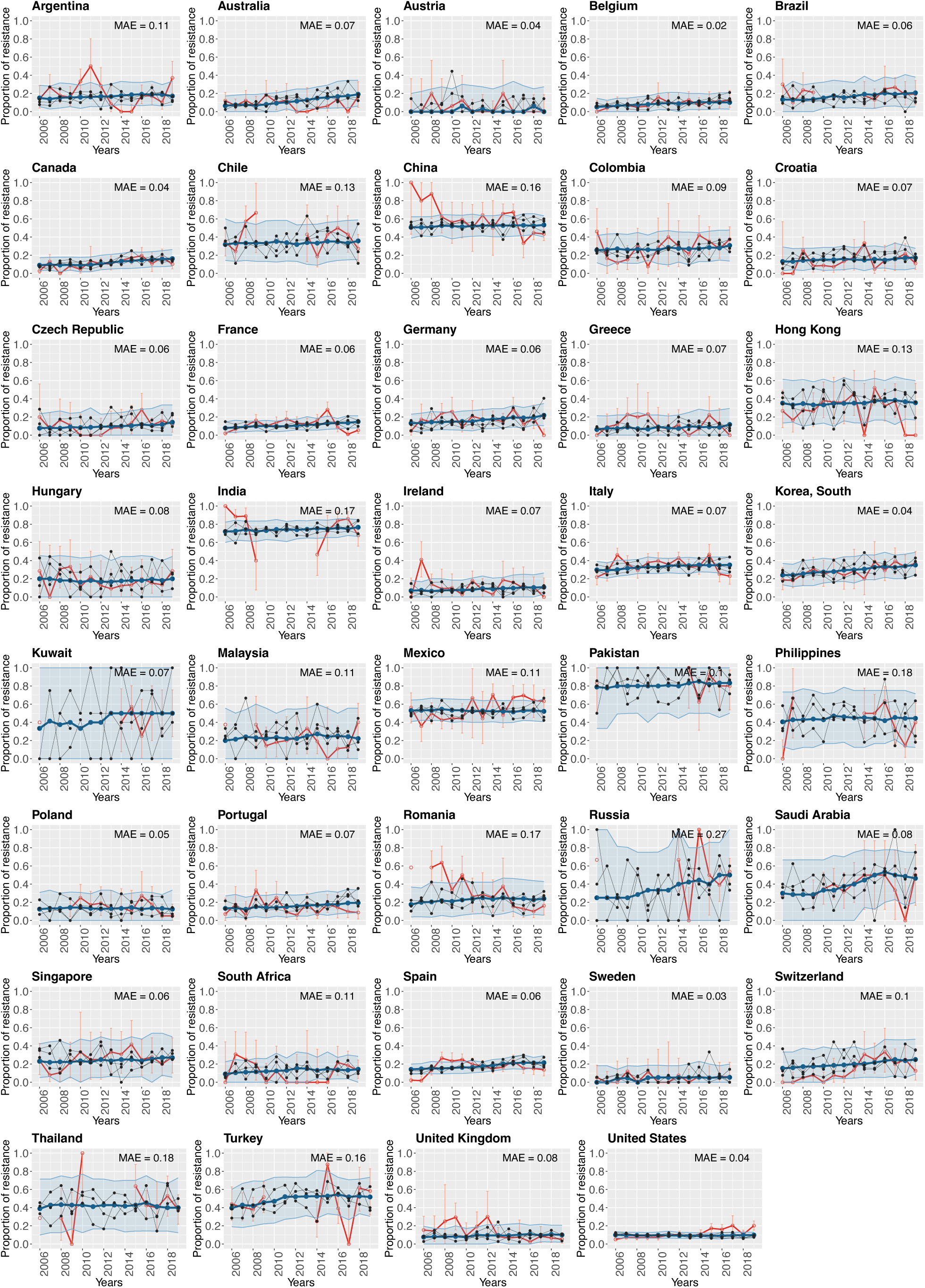
Country-by-country fits of the selected model. In blue, median and prediction intervals from 200 runs using estimated parameters from the selected model (model 6). In dark grey, three randomly selected stochastic runs of the selected model. In red, reported rates of ESBL-production among *E. coli* isolates from bloodstream infections observed in the ATLAS data.^25^ On the top right, associated mean absolute errors (MAE) of observed relative to estimated resistance rates.

### Simulating the hypothetical global emergence of a CR-EC clone

Using the model parameters estimated for ESBL-EC, a simulation study was run to explore expected spatio-temporal patterns and trends of a putative emerging CR-EC clone.

For the four included CR-EC emergence scenarios, annual infection incidence predictions at 5- and 20-years post-emergence are presented in Figure 5. For the “base case” scenario s1, at 5-years post-emergence, incidence was greatest within each index country (matrix diagonal) as well as in nearby countries in the same global region. Conversely, at 20 years, incidence was greatest in countries associated with high estimated transmission rates (*β_i_*) such as Thailand, Philippines, China, and Kuwait, irrespective of the index country. This suggests that dissemination is no longer shaped by international travel but by within-country transmission and selection pressure. For the “fitness cost” scenario s2, dynamics were similar to the base case scenario, but with lower overall incidence due to the assumption of a resistance-associated fitness cost. For the “super-bug” scenario s3, global spread was accelerated, with a greater share of countries having local spread at 5-years, and with high incidence at 20-years, including up to 100 infections per 100,000 inhabitants per year in India and Pakistan. Finally, for the “multi-resistance” scenario s4, the highest incidence countries at 20 years differed from those in scenarios s1 and s3. Notably, Saudi Arabia had among the highest predicted incidence (approximately 50 per 100,000 for scenario s4, compared to <10 per 100,000 for scenarios s1 and s3), likely due to a sharp rise in β-lactams treatment rates observed in this country (Figure 2).

**Figure 5.**
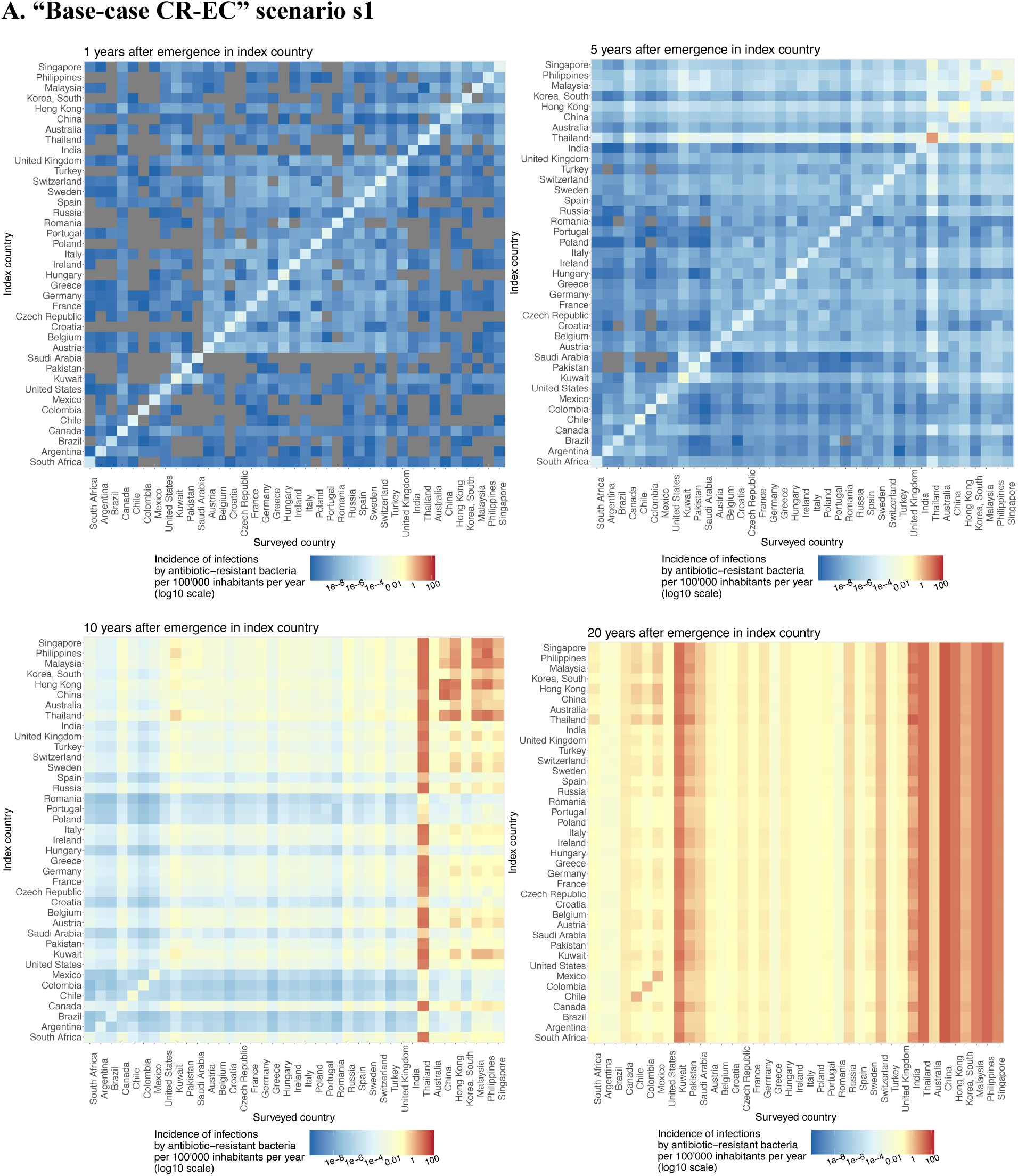
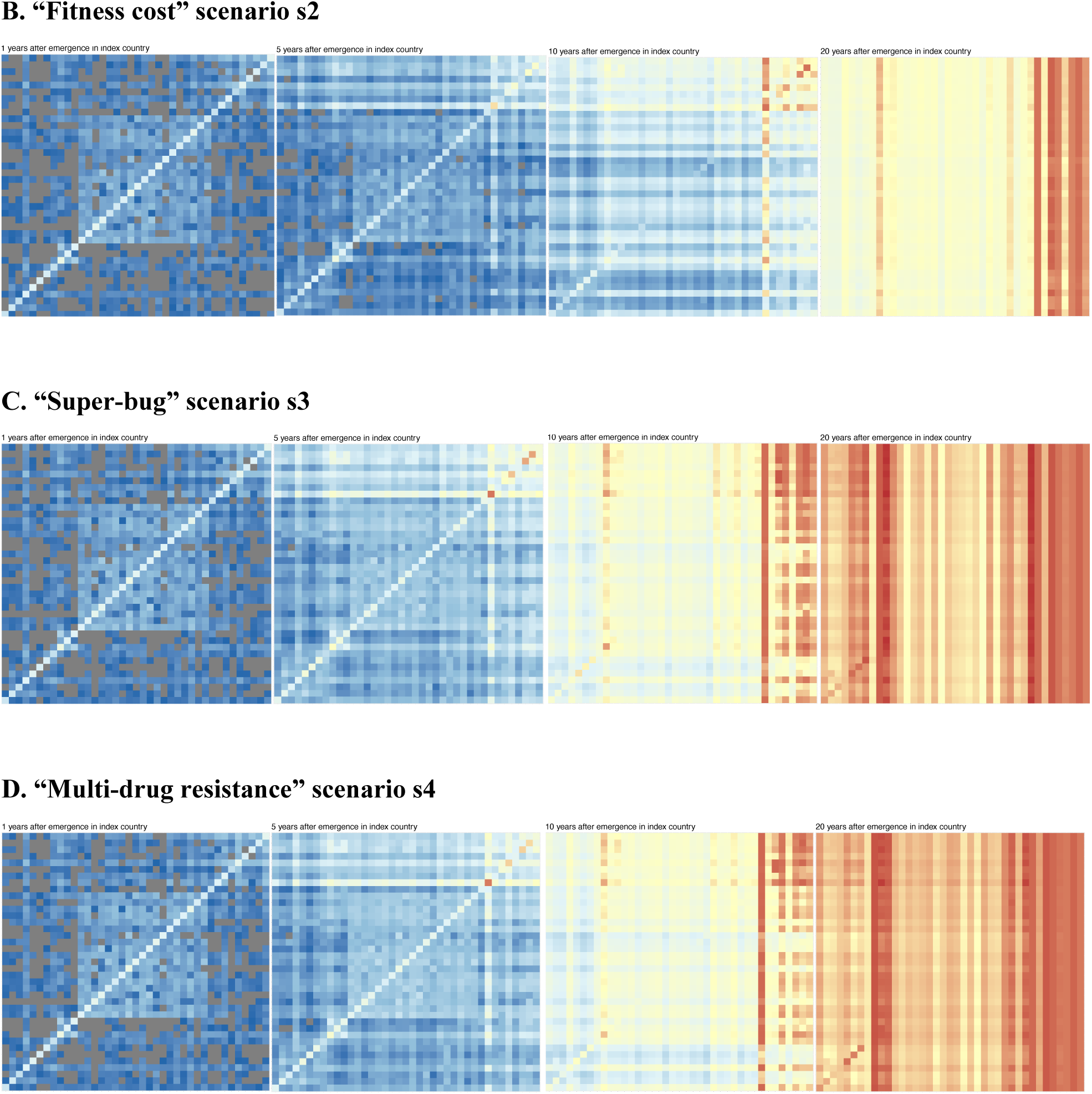
Predicted global dynamics of a hypothetical emergent carbapenem-resistant *E. coli* (CR-EC) clone according to the four scenarios. The figure shows heatmaps of the incidence of CR-EC infections in surveyed countries at 1-, 5-, 10- and 20-years post-emergence in index countries (y-axis). Countries are ordered by global region. Incidence is shown in log10 scale. Grey are zero values indicating no spread in that country at that time. Four scenarios according to the properties of the emerging resistant clone are explored: **(A)** “base-case CR-EC” scenario s1 (β_i_ (s1) = β_i_); **(B)** “fitness cost” scenario s2 (β_i_ (s2) = β_i_×0,9); **(C)** “super-bug” scenario s3 (β_i_ (s3) = β_i_×1,1); **(D)** “multi-drug resistance” scenario s4 (βi (s4) = βi and τ_y,i_ = per capita treatments per day of carbapenems and β-lactams). In all scenarios, the system was initialized with 20 colonized individuals in each index country. In panels B, C and D, countries are ordered as in A.

Finally, we simulated the emergence of CR-EC in each index country with and without further global spread mediated by international travel (Figure 6). Across all scenarios, accounting for CR-EC importation via travel led to greater spread in most countries, including higher incidence at 20 years post-emergence in 32/39 countries for s1, 36/39 for s2, and 31/39 for s3 and s4. Remaining countries with no difference in incidence when allowing importation via travel, such as Thailand, were associated with high local transmission rates.

**Figure 6.**
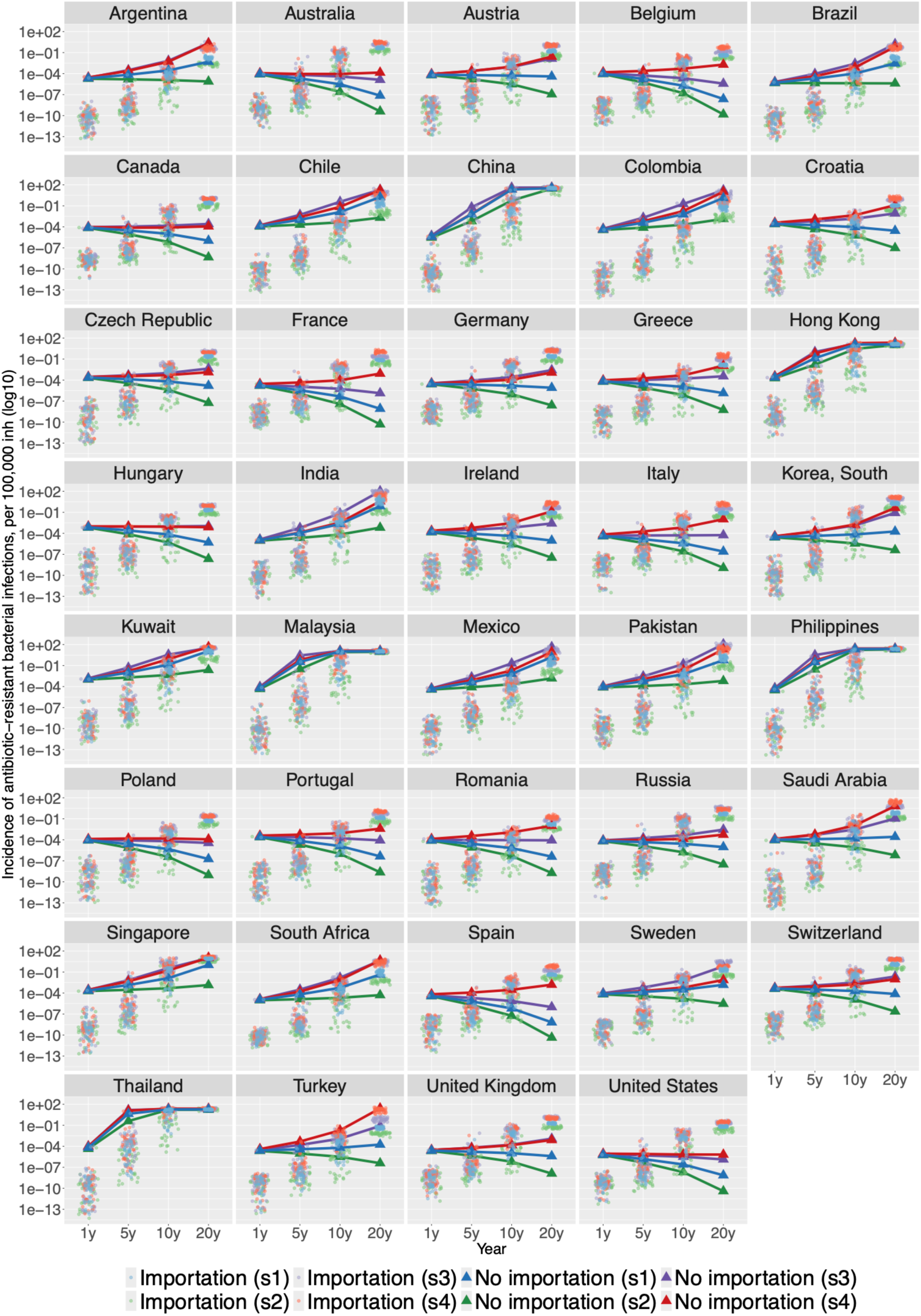
Predicted trends of an emerging carbapenem-resistant *E. coli* clone with and without importation in surveyed countries according to the four scenarios. Trajectories with importation from all possible index country (points) and trajectories without importation (triangles) are shown for the “base-case” scenario s1 in blue (β_i_ (s1) = β_i_), for the “fitness cost” scenario s2 in green (β_i_ (s2) = β_i_×0,9), for the “super-bug” scenario s3 in purple (β_i_ (s3) = β_i_×1,1) and the “multi-drug resistance” scenario s4 in red (βi (s4) = βi and τ_y,i_ = per capita treatments per day of carbapenems and β-lactams). In all scenarios, the system was initialized with 20 colonized individuals in each index country. Index countries are all countries for the “importation” scenario, and only the target country in the “no importation” scenario. Infection incidence on the y-axis is shown in log10 scale. Years on the x-axis corresponds to 1-, 5-, 10- and 20-year post-emergence in the index country.

## Discussion

In this work, we developed a metapopulation compartmental model of antibiotic-resistant *E. coli* transmission, colonization and infection. This study provides, to our knowledge, among the first mechanistic and predictive insights into the spread of antibiotic-resistant *E. coli* at a global scale.^28^ By calibrating the model to available data for 39 countries between 2006 and 2019, we first explored hypotheses explaining observed heterogeneity in the prevalence of ESBL-EC in both colonization and infection globally. We showed that individual antibiotic exposure, but mostly heterogeneous transmission rates across countries, were key to reproduce the observed global infection trends. Using this model calibrated to ESBL-EC data, we further explored several scenarios of the global spread of a hypothetical emergent CR-EC clone. We found that while international travel drove early transmission patterns worldwide, long-term patterns were mostly driven by within-country transmission rates and antibiotic selection pressure. aIn a counterfactual scenario, we showed that predicted long-term antibiotic resistance trends in countries with high local transmission rates can reach high levels even in absence of repeated importation of antibiotic resistance through travel; while in countries with lower transmission levels, continuous importation is needed to maintain transmission and reach fixation.

In our model, antibiotics impacted the transmission of antibiotic-resistant bacteria through two mechanisms: antibiotic-induced microbiome dysbiosis leading to reduced colonization resistance and, in turn, an increased risk of acquiring colonization upon exposure; and a prolonged duration of dominant antibiotic-resistant colonization after treatment cessation.^23^ In fitting our model to data, we estimated a significantly increased risk of acquiring ESBL-EC during treatment. These results are consistent with modeling studies in nosocomial contexts, where antibiotic exposure is more frequent, which have also predicted a significant impact of antibiotic-induced microbiome dysbiosis on ESBL-EC acquisition.^29^ Antibiotic exposure quantities analyzed here were collected from official sales data from retailers or hospitals reported in the IQVIA MIDAS database and do not include donations or sales of antibiotics via parallel markets.^30^ In some countries, these quantities might not be fully representative of the total national antibiotic use, leading to underestimation of the antibiotic consumption and therefore possibly underestimation of the weight of antibiotic selection and overestimation of within-country transmission rates. Nonetheless, the IQVIA MIDAS database represents the most comprehensive global data source available. Due to data limitations, we did not account for heterogeneity in antibiotic use across age classes, settings (hospital versus community) or sub-national regions, which have all been shown to play a role in the geographical distribution of antibiotic resistance.^31^ In the future, models structured for both hospital and community populations in each country should be considered, as Kachalov and colleagues showed that national prevalence of another ESBL-producing Enterobacterales, *K. pneumoniae*, was more strongly impacted by reducing hospital antibiotic consumption than community consumption.^32^

Necessary model simplifications notwithstanding, our results suggest that national-level β-lactam antibiotic consumption is not sufficient to explain all observed heterogeneity in ESBL-EC dynamics, with country-specific transmission rates also playing a critical role. Epidemiologically, such heterogeneity could reflect diverse mechanisms putatively affecting ESBL-EC transmission, such as hygiene, healthcare system organization, environmental and animal exposure or climate.^33,34^ It is important to highlight that within-country transmission parameter estimates are sensitive to the data used for model calibration. First, colonization data were derived from the meta-analysis by Bezabih and colleagues, based on a limited number of mostly cross-sectional studies with small sample sizes.^8^ However, in the absence of systematic longitudinal ESBL-EC colonization monitoring, these data remain the most comprehensive estimates available. Similarly, infection risks estimates are sensitive to *E. coli* bloodstream infections incidence data used in the calibration, which were derived from various studies representing heterogeneous populations and catchment areas (supplementary Table 3). The observed antibiotic resistance trends in infection were derived from the ATLAS monitoring system. However, the representativeness of ATLAS at the country level may be limited. Reporting is conducted by a non-representative selection of hospitals, potentially introducing bias in observed resistance rates. Moreover, sampled isolates originate predominantly from severe infections, which may exhibit higher resistance rates due to hospitalization following failed community treatment. Given our model’s assumption that resistance in colonization is proportional to resistance in infection, this could lead to overestimation of global ABR rates and bias in the estimation of within-country transmission rates in colonization. Finally, small sample sizes for certain country–year observations contribute to high uncertainty in resistance rate estimates. Nevertheless, previous studies indicate that, while ATLAS reports higher resistance frequencies than publicly available surveillance programs (like the ECDC), the overall dynamics of resistance trends may still be accurately captured.^6,35^ Consequently, although absolute values of within-country transmission rates may be overestimated, our comparative analysis of global inter-country dynamics remains valid. In addition, frequent acquisition of plasmids and antibiotic resistance genes has been observed in *E. coli*.^36^ Our model did not account for these gene transfers within the microbiota, nor environmental or zoonotic acquisition, relying solely on human-to-human transmission of resistant bacteria. Adding such mechanisms to models will require robust experimental and environmental data to inform model parameterization and discriminate between the various putative sources of antibiotic resistance acquisition in humans. Finally, there is evidence that demographic shifts such as changes in population size or ageing communities will lead to an increase in antibiotic-resistant infections incidence.^1^ Better capturing demography dynamics into models, which was not done in our study, might thus improve the predictions of future antibiotic resistance trends at the global level.

Several studies have reported a high prevalence of colonization by ESBL-producing Enterobacterales in returning travelers, with prevalence varying depending on the destination.^17^ However, travelers’ contributions to the global burden of ESBL-EC infections has not been quantified. In this study, we found that the role of international travel in shaping global antibiotic resistance levels and spread was both time- and country-dependent. Our simulations showed that travel played a significant role on the spread of a hypothetical CR-EC clone shortly after its emergence. But its role faded over time, at different rates depending on the clone’s transmission characteristics and countries’ selective pressure. At 20-years post-emergence, the predominant factors shaping incidence and resistance globally were the within-country human-to-human transmission rate and antibiotic consumption. Results from multivariable sensitivity analyses – carried out for two different model initializations, “endemic” and “emergent” (supplementary Table 4, Figures 4 and 5) – corroborated such findings. However, in countries with lower within-country transmission rates, travel was necessary to maintain transmission and to reach antibiotic resistance endemic levels (like the United States for example). It is important to stress that contacts during travel in the destination country was assumed to be homogeneous in terms of risk, which might not be true in case of forced migration, war displacements or medical tourism (e.g. between-country transfers of sick or hospitalized patients). Moreover, in order to simplify the model, we focused on the risk associated with importation of antibiotic-resistant bacteria from returning travelers and neglected individuals traveling from other countries possibly carrying and exporting resistance. Such one-direction hypothesis might lead to an underestimation of travelers’ role in disseminating antibiotic-resistant clones globally.

In conclusion, we have developed a mechanistic model that reproduces global trends in ESBL-EC transmission, colonization and infection from 2006 to 2019. Country-specific variation in human-to-human transmission risk was identified as the key driver of global heterogeneity in ESBL-EC epidemiology, along with individual β-lactam exposure. Simulations were then used to anticipate global dissemination risks associated with the emergence of a novel CR-EC clone, highlighting the key role of travel in shaping early global spread before the local establishment of endemic transmission. Conversely, these results reinforce that once a clone has become endemic, controlling local transmission risk and antibiotic selection, for instance through robust sanitation infrastructure and antibiotic use policies, are critical to minimize infection burden. Overall, our work highlights how mathematical models make it possible to analyze spatiotemporal dynamics and anticipate the possible spread of new emerging superbugs. While specific to *E. coli*, this approach could be adapted to evaluate similar questions for other bacterial species. However, models are mainly limited by the scarcity of high-quality data, especially comparable prevalence of sensitive and resistant bacterial colonization across countries. Future efforts on data collection will be required to better inform models and further use them to support public health decision making for mitigating the impact of antibiotic resistance on human health.

## Data Availability

All data are available online at https://atlas-surveillance.com/login. All data produced in the present study are available upon reasonable request to the authors.

