## Appendix for "Global dynamics of antibiotic resistance: a modeling study of the spread of endemic and emerging resistance in *E. coli*"

#### Supplementary Methods and Results.


1. Model description. 2
2. Data used for input parameters or calibration. 9
3. Multivariable sensitivity analyses. 16
4. Procedure for model calibration and results for model comparison. 22
5. Country-independent model calibration. 23
6. MCMC results for selected model (model 6). 24

#### 1. Model description.

***Equation S1***

##### Model equations

The transmission dynamics of antibiotic-resistant *E. coli* within a given country *i* is modeled by a system of ordinary differential equations described below. The model is implemented in R (version 4.2.1) using the *odin* package. (FitzJohn R. ODE Generation and Integration [R package odin version 1.2.5] - 10.32614/CRAN.package.odin)

**Notation**

Country $i$ with $i = 1, \ldots, n$ ($n$ = number of countries analyzed)

Country $j$ with $j = 1, \ldots, n$

Year y with y = 2006, …, 2019

**Ordinary Differential Equations for country *i***

$$\frac{d\boldsymbol{CS_i}}{dt} = -CS_i \times [\sum_{j=1}^{n} \Delta_{y,ij} \times \boldsymbol{\lambda_j}] + CSr_i \times \gamma - CS_i \times \tau_{i,y} + CS^A{}_i \times \frac{1}{\alpha}$$

$$\frac{d\boldsymbol{CSr_i}}{dt} = CS_i \times [\sum_{j=1}^{n} \Delta_{y,ij} \times \boldsymbol{\lambda_j}] - CSr_i \times \gamma - CSr_i \times \tau_{i,y} + CsR_i \times \frac{1}{\alpha'}$$

$$\frac{d\boldsymbol{CsR_i}}{dt} = -CsR_i \times \frac{1}{\alpha'} + CsR^A{}_i \times \frac{1}{\alpha} - CsR_i \times \tau_{i,y}$$

$$\frac{d\boldsymbol{CS^A{}_i}}{dt} = -CS^A{}_i \times \lambda_{ant\, i} + CS_i \times \tau_{i,y} - CS^A{}_i \times \frac{1}{\alpha}$$

$$\frac{d\boldsymbol{CsR^A{}_i}}{dt} = CS^A{}_i \times \lambda_{ant\, i} + CSr_i \times \tau_{i,y} + CsR_i \times \tau_{i,y} - CsR^A{}_i \times \frac{1}{\alpha}$$

With the following compartments: $\boldsymbol{CS_i}$ individuals colonized with antibiotic-Sensitive strain; $\boldsymbol{CSr_i}$ individuals colonized with antibiotic-Sensitive strain and sub-dominant antibiotic-Resistant strain; $\boldsymbol{CsR_i}$ individuals colonized with antibiotic-Sensitive strain and dominant antibiotic-Resistant strain; $\boldsymbol{CS^A{}_i}$ individuals colonized with antibiotic-Sensitive strain and exposed to antibiotics; and $\boldsymbol{CsR^A{}_i}$ individuals colonized with antibiotic-Sensitive strain and dominant antibiotic-Resistant strain and exposed to antibiotics.

Compartments are all time dependent, and evaluated in days; but are noted without the time dependent notation *(t)* for clarity.

**Forces of colonization for country i**

Force of colonization $\lambda_i$ :

$$\lambda_i = \frac{(CSr_i + CsR_i + CsR^A{}_i)}{N_i} \times \beta_i$$

Force of colonization under antibiotic treatment $\lambda_{ant\,i}$ :

$$\lambda_{ant\,i} = \lambda_i \times \phi_{ant}$$

**Mobility matrix, $\Delta_y$, size $n \times n$**

$$\Delta_y = \begin{bmatrix} 1 & \dots & \frac{Number\ of\ travelers\ from\ country\ 1 \to n}{Population\ size\ country\ 1}/365 \\ \dots & 1 & \dots \\ \frac{Number\ of\ travelers\ from\ country\ n \to 1}{Population\ size\ country\ n}/365 & \dots & 1 \end{bmatrix}$$

where $\Delta_{y,ij}$ is the proportion of individuals of country *i* going from country *i* to country j each day during year *y*

**Total population for country *i* at time $t_0$**

$$N_i = CSr_i + CsR_i + CsR^A{}_i + CS_i + CS^A{}_i$$

The size of the population for each country was kept constant and matched to World Bank average population sizes over the study period (supplementary Table S2).

#### Model outcomes

Numerical integration of the ODEs described above is used to calculate various model outcomes for each country *i* and each year *y* over a 14-year period (2006-2019), after model equilibrium has been reached for the year 2006. The ODEs are solved by numerical integration using the lsoda method, from the ode function in the *deSolve* package. (Soetaert K. Solvers for Initial Value Problems of Differential Equations ('ODE', 'DAE', 'DDE') [R package deSolve version 1.40]. - 10.32614/CRAN.package.deSolve)

**Infections in country *i*, for period t to t+1 (in days)**

The number of new infections with antibiotic-sensitive bacteria is calculated as:

$$IS_i = \int_t^{t+1} \sigma_{S\,i} \times [CS_i + CSr_i \times k]\ dt$$

The number of new infections with antibiotic-resistant bacteria is calculated as:

$$IR_i = \int_t^{t+1} \sigma_{R\,i} \times [CSr_i \times (1-k) + CsR_i + CsR^A{}_i]\ dt$$

with $\sigma_{R\,i} = a_i \times \sigma_{S\,i}$

**Incidence per 100,000 inhabitants of total infections in country *i***

$$inc_{infection\,i} = \frac{IR_i + IS_i}{N_i} \times 100000$$

**Proportion of antibiotic resistance in infection (resistance rate)**

$$RR_{infection\ i} = \frac{IR_i}{IR_i + IS_i} = \frac{IR_i}{I_i}$$

**Proportion of antibiotic resistance in colonization (resistance rate)**

$$RR_{colonized\ i} = \frac{CSr_i + CsR_i + CsR^A{}_i}{CSr_i + CsR_i + CsR^A{}_i + CS_i + CS^A{}_i} = \frac{CSr_i + CsR_i + CsR^A{}_i}{N_i}$$

***Equation S2***

#### Stochastic observation model

The number of infections with antibiotic-resistant bacteria reported by the ATLAS surveillance system for country *i* and year *y* $r_{i,y}$ is modeled as follows:

$$r_{i,y} \sim Bin(IR_{i,y} , \frac{tot_i}{tot\_est_i})$$

- $IR_{i,y}$ is the number of antibiotic-resistant infections predicted by the model
- $tot_i$ is the total number of infections reported by the surveillance system for country *i*, cumulated over years
- $tot\_est_i$ is the total number of infections estimated for country *i*, based on the calculation $tot\_est_i$ = (incidence data for country $i \times N_i$ ) / 100000, cumulated over years

***Equation S3***

#### Likelihood

The expression for the likelihood *L* is as follows:

$$\mathcal{L}(data|parameters)$$

$$= \mathcal{L}(RR_{colonized}) \times \mathcal{L}(inc_{infection}) \times \mathcal{L}(resistant\ infections\ ) \times \mathcal{L}(sensitive\ infections)$$

$$= \prod_i \prod_y P(\overline{RR}^{obs}_{colonized\ i,y} | \overline{RR}_{colonized\ i,y}) \times P(inc^{obs}_{infection\ i,y} | inc_{infection\ i,y})$$

$$\times P(r_{i,y}\ \ tot_{i,y} | IR_{i,y}) \times \mathrm{P}(s_{i,y}\ \ tot_{i,y} | IS_{i,y})$$

The number of infections with antibiotic-resistant bacteria reported in ATLAS $r_{i,y}$ by country *i* and year *y* is assumed to be distributed according to a binomial distribution, such that:

$$P(data|parameters) = \prod_i \prod_y P(r_{i,y} , tot_{i,y} | IR_{i,y})$$

$$r_{i,y} \sim Bin(IR_{i,y}\ ,\ \theta_{i,y} = \frac{tot_{i,y}}{I_{i,y}})$$

The number of infections with antibiotic-sensitive bacteria reported in ATLAS $s_{i,y}$ by country *i* and year *y* is assumed to be distributed according to a binomial distribution, such that:

$$P(data|parameters) = \prod_{i}\prod_{y} P(s_{i,y}\ ,\ tot_{i,y} \middle| IS_{i,y})$$

$$s_{i,y} \sim Bin(IS_{i,y}\ ,\ \theta_{i,y} = \frac{tot_{i,y}}{I_{i,y}})$$

The incidence of *E. coli* bloodstream infections $inc_{infection\ i,y}^{obs}$ by region (used for each country *i*) and year *y* is assumed to be distributed according to a Poisson distribution, such that:

$$P(data|parameters) = \prod_{i}\prod_{y} P(inc_{infection\ i,y}^{obs} \middle| inc_{infection\ i,y})$$

$$inc_{infection\ i,y}^{obs} \sim Pois\left(inc_{infection\ i,y}\right)$$

$$\text{With} \begin{cases} if\ y = year_{literature};\ inc_{infection}^{obs} \neq NA \\ if\ y\ \neq year_{literature};\ inc_{infection}^{obs} = NA \end{cases}$$

The proportion of individuals asymptomatically colonized by antibiotic-resistant bacteria $RR_{colonized\ i,y}^{obs}$ by country *i* and year *y* is assumed to be normally distributed, such that:

$$P(data|parameters) = \prod_{i}\prod_{y} P(RR_{colonized\ i,y}^{obs} \middle| RR_{colonized\ i,y})$$

$$RR_{colonized\ i,y}^{obs}\ \sim N(RR_{colonized\ i,y}\ , var)$$

$$\text{With} \begin{cases} if\ y = year_{literature};\ RR_{colonized}^{obs} \neq NA \\ if\ y\ \neq year_{literature};\ RR_{colonized}^{obs} = NA \end{cases}$$

#### Model parametrization

Parameters are listed below in supplementary Table 1.

**Antibiotic consumption data.** Annual antibiotic consumption data for each country over the period 2006-2019 are taken from the MIDAS IQVIA database. (1) Antibiotic use is reported in kilograms and represents molecules sold both in hospitals and in the community. We convert these data into treatment rates per individual, per day and per country. We consider here that 20 grams of β-lactam antibiotic molecule over 8 days represents standard antibiotic therapy for an individual. (2) For the selection of ESBL-producing *E. coli*, the model uses national consumption of β-lactams, including broad-spectrum penicillins, monobactams and cephalosporins.

**Travel data.** The mobility matrix is parameterized using data from the Knowledge Center for Migration and Demography. (3) These data represent the estimated cumulative annual number of travelers between two countries, for each year between 2011 and 2016. For the years not available (2006-2010 and 2017-2019), a linear regression extrapolation is performed. Statistically significant linear trends over 2011-2016 are maintained. If no trend is observed over 2011-2016, a constant trend is applied. The final estimated mobility matrix is shown in supplementary Figure 1.

**Demographic data.** Population size is defined by the number of inhabitants per country studied, obtained from World Bank values averaged over the period 2006-2019. (4)

**Initial values for MCMC algorithm.** The initial value of $\sigma_{S\,i}$ used in the MCMC algorithm is constructed as follows. We assume that the entire population of a country is colonized by at least one *E. coli* strain. Neglecting colonized individuals of antibiotic-resistant strains in a low-prevalence country, such as the United Kingdom, we derive a probability of infection by an *E. coli* strain from the annual incidence of bloodstream infections in that country: 60/100,000/365 days = $1.6\text{x}10^{-6}$ day$^{-1}$. This number is then used as initial value in the MCMC procedure for the $\sigma_{S\,i}$ parameter, for all countries.

***Table S1***

**Parameters interpretation, units, and sources.** MCMC: Markov Chain Monte Carlo

| Parameter | Interpretation | Parameter derivation | Values & units | Source |
|---|---|---|---|---|
| $\gamma$ | Natural clearance rate for ESBL-producing *E. coli.* | Direct from source | 1/120 day$^{-1}$ | van den Bunt, 2020 (5) |
| $\alpha$ | Duration of antibiotic treatment. | Direct from source | 8 days | Average based on Esposito, 2012 (6) |
| $\alpha'$ | Duration of microbiome dysbiosis (transient period of | Direct from source | 60 days | Palleja, 2018 (7) |

| | | | | |
|---|---|---|---|---|
| | imbalance after antibiotic exposure). | | | |
| $k$ | Proportion of co-colonized individuals (dominant sensitive strain and subdominant resistant strain) who develops an infection by the sensitive strain. | Assumed | 0.80 | - |
| $\tau_{y,i}$ | Individual antibiotic exposure rate by year *y* and country *i*. | Calculated from data | $day^{-1}$ | Data from MIDAS IQVIA (1), see below for details |
| $\Delta_y$ | Mobility matrix for year *y* representing the percentage of inhabitants of country *i* traveling to country *j*. | Calculated from data | % | Data from KCMD (3), see below for details |
| $N_i$ | Population size by country *i*. | Calculated from data | Number of inhabitants. | World Bank data (4), see details above |
| $\phi_{ant}$ | Excess risk of ESBL-producing *E. coli* acquisition for individuals exposed to antibiotics. | Bayesian inference | Initial value (MCMC): 10<br><br>A priori distribution (MCMC): Uniform [min = 0; max = 50]. | / |
| $\beta_i$ | Transmission rate for ESBL-producing *E. coli.* | Bayesian inference | Initial value (MCMC): $9x10^{-3}$ $day^{-1}$<br><br>A priori distribution (MCMC): Uniform [min = $1x10^{-6}$; max = $1x10^{-1}$] | / |
| $\sigma_{S\ i}$ | Rate of progression from colonization to infection for individuals colonized by antibiotic-sensitive *E. coli.* | Bayesian inference | Initial value (MCMC): $1.6x10^{-6}$ $day^{-1}$<br><br>A priori distribution (MCMC): Uniform [min = $1x10^{-6}$ ; max = $2x10^{-6}$] | See details above |
| $a_i$ | Colonized-to-infected progression rate ratio for antibiotic-resistant vs sensitive strains, a : $\sigma_{R\ i} = a_i \times \sigma_{S\ i}$. | Bayesian inference | Initial value (MCMC): 20<br><br>A priori distribution (MCMC): Uniform [min = 0; max = 100]. | / |

*(3)* **Knowledge Centre on Migration and Demography (KCMD) Data Portal.** https://migration-demography-tools.jrc.ec.europa.eu/data-hub/

*(4)* **World Bank Data**. https://data.worldbank.org

*(5)* van den Bunt G, Fluit AC, Bootsma MCJ, *et al.* **Dynamics of Intestinal Carriage of Extended Spectrum Beta-lactamase-Producing Enterobacteriaceae in the Dutch General Population, 2014-2016**. *Clin Infect Dis* 2020; **71**: 1847–55.

*(6)* Esposito S, Esposito I, Leone S. **Considerations of antibiotic therapy duration in community- and hospital-acquired bacterial infections.** *J Antimicrob Chemother* 2012; **67**: 2570–5.

*(7)* Palleja A, Mikkelsen KH, Forslund SK, *et al.* **Recovery of gut microbiota of healthy adults following antibiotic exposure.** *Nat Microbiol* 2018; **3**: 1255–65.

#### 2. Data used for input parameters or calibration.

##### Demographic data

***Table S2. Demographic data for the countries included in the modeling analysis.***

Countries, associated regions, and average population size over 2006-2019 from World Bank Data. (https://data.worldbank.org)

| Country | World's region | Population |
|---|---|---|
| Argentina | Americas | 41611127 |
| Australia | W-Pacific | 22603338 |
| Austria | Europe | 8483405 |
| Belgium | Europe | 10992563 |
| Brazil | Americas | 198103180 |
| Canada | Americas | 34568887 |
| Chile | Americas | 17377114 |
| China | W-Pacific | 1347845000 |
| Colombia | Americas | 45979844 |
| Croatia | Europe | 4240407 |
| Czech Repubic | Europe | 10456372 |
| France | Europe | 65319205 |
| Germany | Europe | 81904851 |
| Greece | Europe | 10945360 |
| Hong Kong | W-Pacific | 7120812 |
| Hungary | Europe | 9939024 |
| India | SE-Asia | 1253967752 |
| Ireland | Europe | 4563900 |
| Italy | Europe | 59499551 |
| Korea, South | W-Pacific | 49970506 |
| Kuwait | E-Mediterranea | 3234032 |
| Malaysia | W-Pacific | 28723524 |
| Mexico | Americas | 116281881 |
| Pakistan | E-Mediterranea | 185770110 |
| Philippines | W-Pacific | 96468760 |
| Poland | Europe | 38061559 |
| Portugal | Europe | 10451237 |

| | | |
|---|---|---|
| Romania | Europe | 20252057 |
| Russia | Europe | 143568422 |
| Saudi Arabia | E-Mediterranea | 28762020 |
| Singapore | W-Pacific | 5111743 |
| South Africa | Africa | 52641040 |
| Spain | Europe | 45944154 |
| Sweden | Europe | 9540856 |
| Switzerland | Europe | 7966387 |
| Thailand | SE-Asia | 67555395 |
| Turkey | Europe | 74611235 |
| United Kingdom | Europe | 63448690 |
| United States | Americas | 311955738 |

#### Mobility data

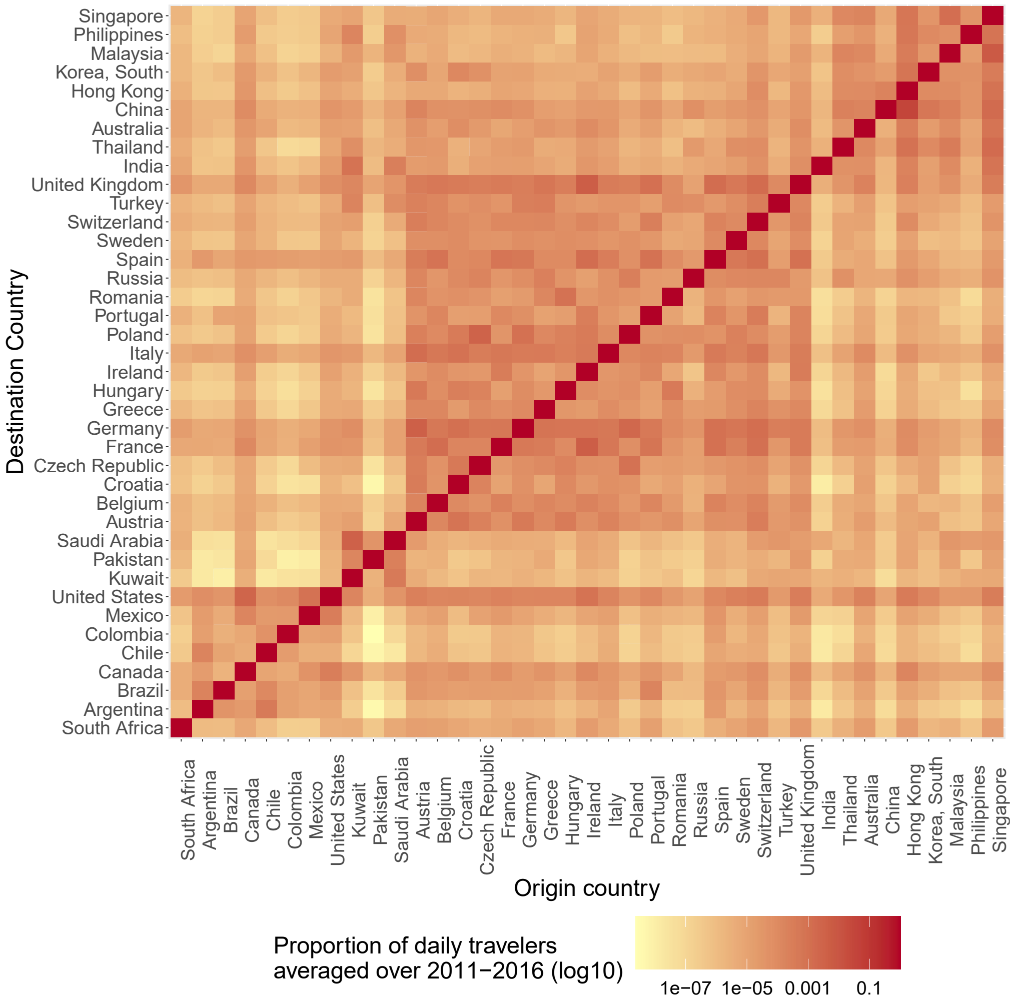


***Figure S1. 2011-2016 averaged mobility matrix Δ.***

Yearly data from KCMD (Knowledge Centre on Migration and Demography (KCMD) Data Portal. https://migration-demography-tools.jrc.ec.europa.eu/data-hub/). For the years not available (2006-2010 and 2017-2019), a linear regression extrapolation is performed. Statistically significant linear trends over 2011-2016 are maintained. If no trend is observed over 2011-2016, we apply a constant trend.

#### Data on asymptomatic colonization by ESBL-producing *E. coli*

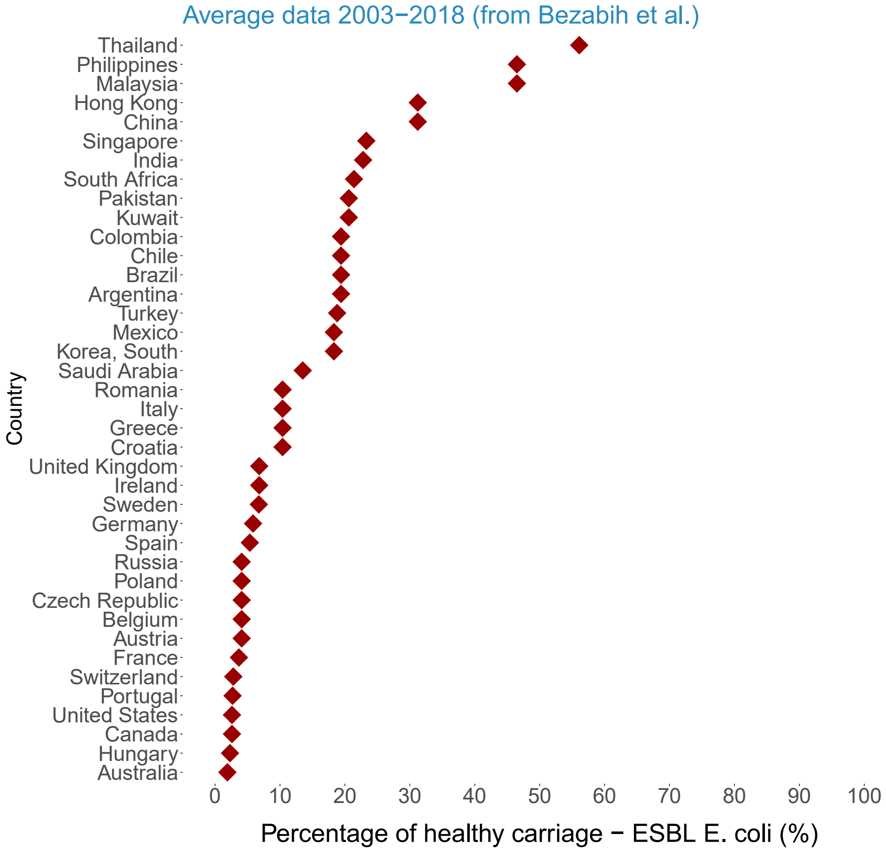


***Figure S2. Proportion of healthy colonization by ESBL-producing E. coli.***

Data by country, averaged over 2003-2018. Data from "Bezabih YM, Sabiiti W, Alamneh E, *et al.* The global prevalence and trend of human intestinal carriage of ESBL-producing Escherichia coli in the community. *J Antimicrob Chemother* 2021; **76**: 22–9."

#### Incidence of infections

***Table S3. Yearly incidence of E. coli bloodstream infections.***

Table with values from the literature for the yearly incidence of *E. coli* bloodstream infections.

| | $inc_{infection}^{obs}$ |
|---|---|
| SE Asia | Thailand – 2010<br>**37/100,000/year** (1) |
| W Pacific | Australia – 2008<br>**50/100,000/year** (2) |
| Africa | Malawi – 2016<br>**20/100,000/year** (3) |
| E Mediterranean | Israel – 2018<br>**90/100,000/year** (4) |
| Americas | Canada – 2016<br>**50/100,000/year** (5) |
| Europe | England – 2016<br>**60/100,000/year** (6) |

*(6)* **Quantifying where human acquisition of antibiotic resistance occurs: a mathematical modelling study.** Knight 2018, BMC

##### Antibiotic consumption data

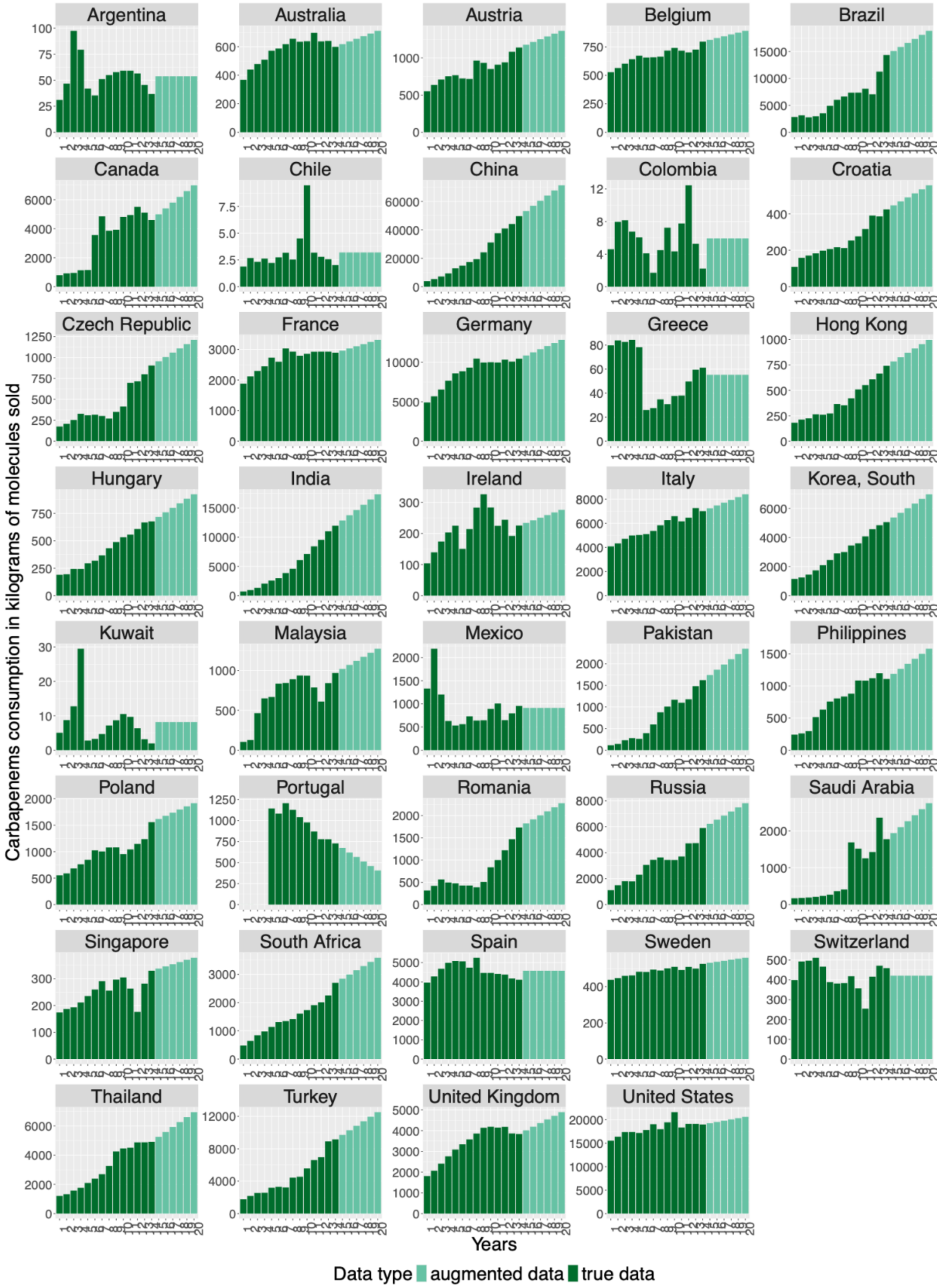

***Figure S3. Carbapenem consumption (observed and augmented data).***

Annual carbapenem consumption in kilograms of molecule sold, by country analyzed. Data augmentation is carried out (pale green) from reported data (dark green) either by maintaining linear trends that are statistically significant over the 2006-2019 period, or by applying constant trends when no significant trend is observed over 2006-2019 (using t-tests for determining significance).

#### 3. Multivariable sensitivity analyses.

##### Procedure

A multivariable sensitivity analysis was performed to quantify the impact of uncertainty around the model input parameters on two model outcomes: the rate of antibiotic resistance in colonized individuals per country for one year (**criteria 1**) and the number of antibiotic-resistant infections per country for one year (**criteria 2**). We show results for a single country, called country A, here Brazil. The parameters included in the sensitivity analysis were different for criteria 1 and 2 (supplementary Table 2). The sensitivity analysis is carried out both for an "**endemic**" model initialization, where the criteria are measured at equilibrium at the end of year 1, and for an "**emergent**" model initialization, where the criteria are measured at the end of year 1 after the introduction of 20 colonized individuals into the CSr compartment of country A at time t=0 day. Parameters are sampled from the hypothetical input parameter distributions (supplementary Table 2) using the Latin hypercubic sampling (LHS) method. N = 200 parameter combinations are generated. Then, the model is simulated N times using these N combinations; and the two criteria of interest are evaluated. Partial rank correlation coefficients (PRCC) are then used to quantify and rank the impact of each parameter on the two chosen criteria (*epiR* package on the R software). (Stenvenson M. epiR: Tools for the Analysis of Epidemiological Data [R package epiR version 2.0.81] 10.32614/CRAN.package.epiR)

##### Results

We found that in the "endemic" case, model outcomes such as antibiotic resistance rates in colonized individuals were strongly sensitive to input parameters β and γ, positively for β and inversely for γ, but not to input parameters linked to travel. Conversely, in the "emergence" case, antibiotic resistance rates in colonized individuals, while still sensitive to β and γ, but also to $\sigma_{Si}$, *a*, and *k*, were strongly sensitive to travel parameters.

*Table S4*

**Hypothetical distributions of model input parameters, used for the LHS sampling.** Example of parameters for country A.

| Parameter | Units | Criteria tested | Hypothetical distribution |
|---|---|---|---|
| $\boldsymbol{\beta_{i=A}}$ | $Day^{-1}$ | Criteria 1 & 2 | Normal [mean = 0.008; standard deviation = 0.002] |
| $\boldsymbol{\sigma_{S\,i=A}}$ | $Day^{-1}$ | Criteria 2 | Normal [mean = $1.3x10^{-6}$ ; standard deviation = $4x10^{-7}$] |
| $\boldsymbol{a_{i=A}}$ | - | Criteria 2 | Uniform [min = 1; max = 100] |
| $\boldsymbol{\phi_{ant}}$ | - | Criteria 1 & 2 | Uniform [min = 2 ; max = 20] |
| $\boldsymbol{\gamma}$ | $Day^{-1}$ | Criteria 1 & 2 | Normal [mean = 1/120; standard deviation = 1/800]. |
| $\boldsymbol{\alpha}$ | Days | Criteria 1 & 2 | Uniform [min = 6 ; max = 10] |
| $\boldsymbol{\alpha'}$ | Days | Criteria 1 & 2 | Uniform [min = 15 ; max = 90] |
| $\boldsymbol{k}$ | - | Criteria 2 | Uniform [min = 0.5; max = 1] |
| $\boldsymbol{\tau_{y=1,i=A}}$ | $Day^{-1}$ | Criteria 1 & 2 | Normal [mean = data(A); standard deviation = data(A)/10] |
| $\boldsymbol{\Delta_{y=1,\ i=A\rightarrow j}}$ | % | Criteria 1 & 2 | Uniform [min = 0 ; max = $5x10^{-4}$] |
| $\boldsymbol{\Delta_{y=1,\ i\rightarrow j=A}}$ | % | Criteria 1 & 2 | Uniform [min = 0 ; max = $5x10^{-4}$] |

#### (A) Criteria 1 (antibiotic-resistance rate) evaluated for country A = Brazil

**Endemic case**

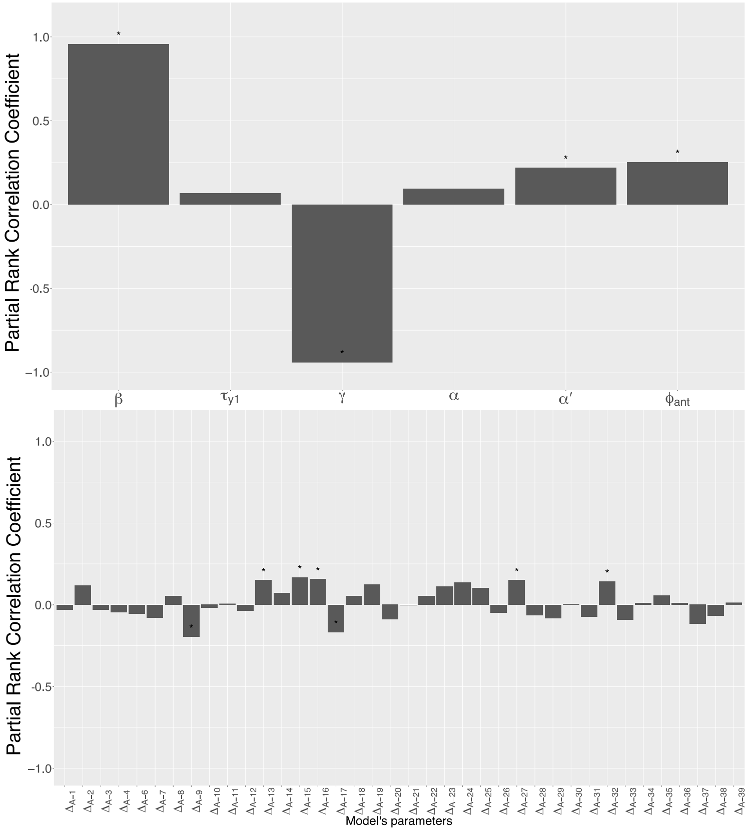


**Emergence case**

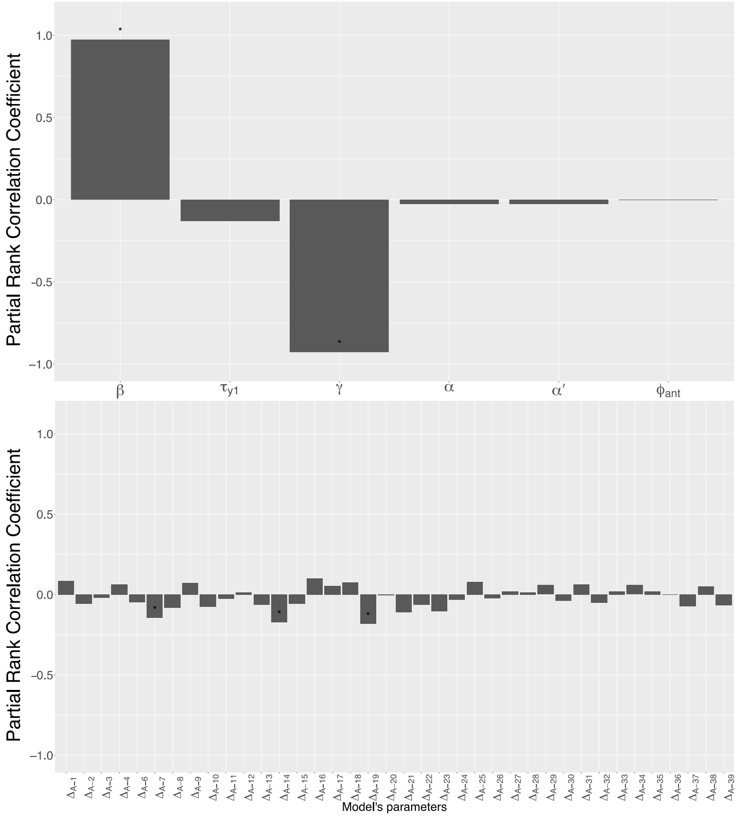

#### (B) Criteria 1 (antibiotic-resistance rate) evaluated for country B

##### Endemic case

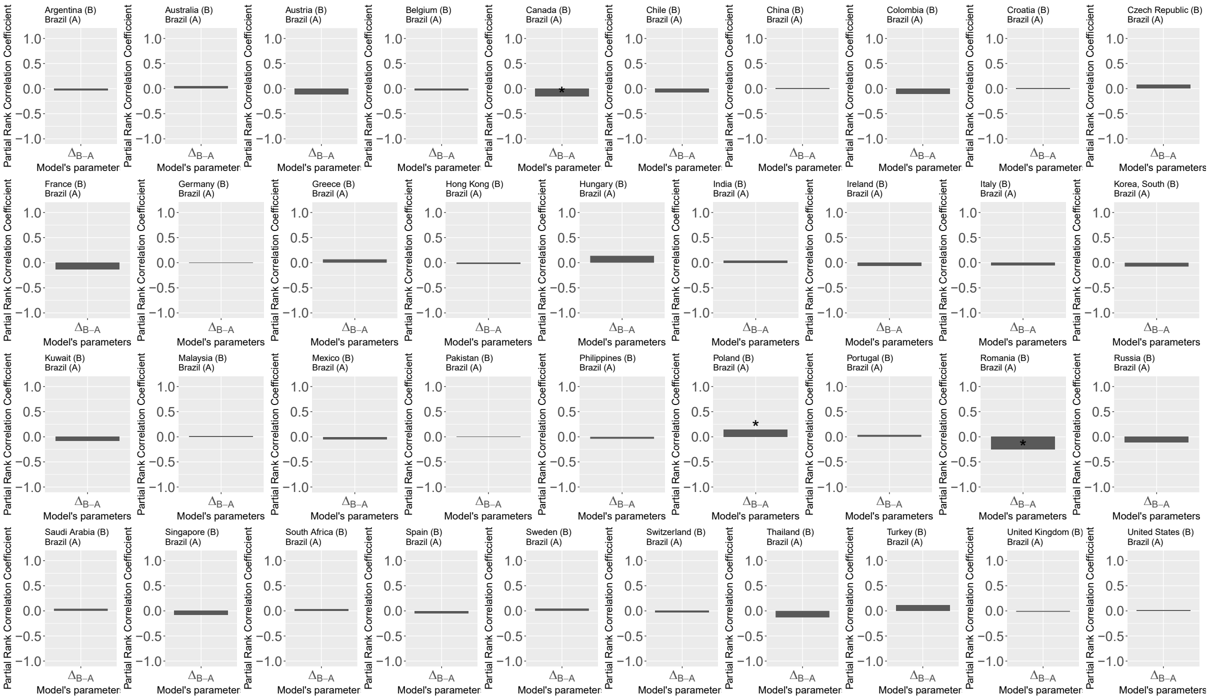


##### Emergence case

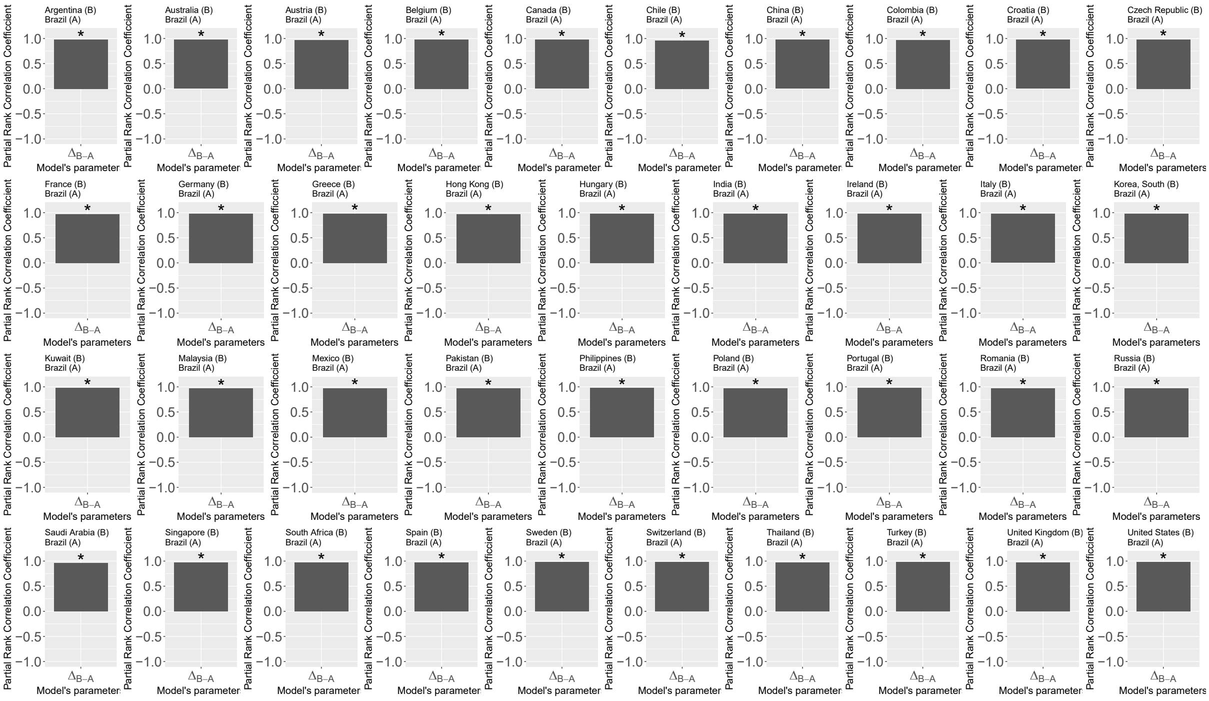


***Figure S4. Sensitivity analysis results: impacts of input parameter uncertainty on the proportion of colonized individuals carrying antibiotic-resistant bacteria (resistance rate in colonization).***

**Partial rank correlation coefficient (PRCC), for the proportion of colonized individuals (criteria 1) for one country and one year – country A: Brazil. (A):** PRCC between model parameters associated to country A (within-country parameters and A-to-B travel parameters) and criteria 1 evaluated for country A. **(B):** PRCC between B-to-A travel parameters and criteria 1 evaluated for country B after introduction in country A. * indicates a significant association (5% threshold).

#### (A) Criteria 2 (number of antibiotic-resistant infections) evaluated for country A = Brazil

**Endemic case**

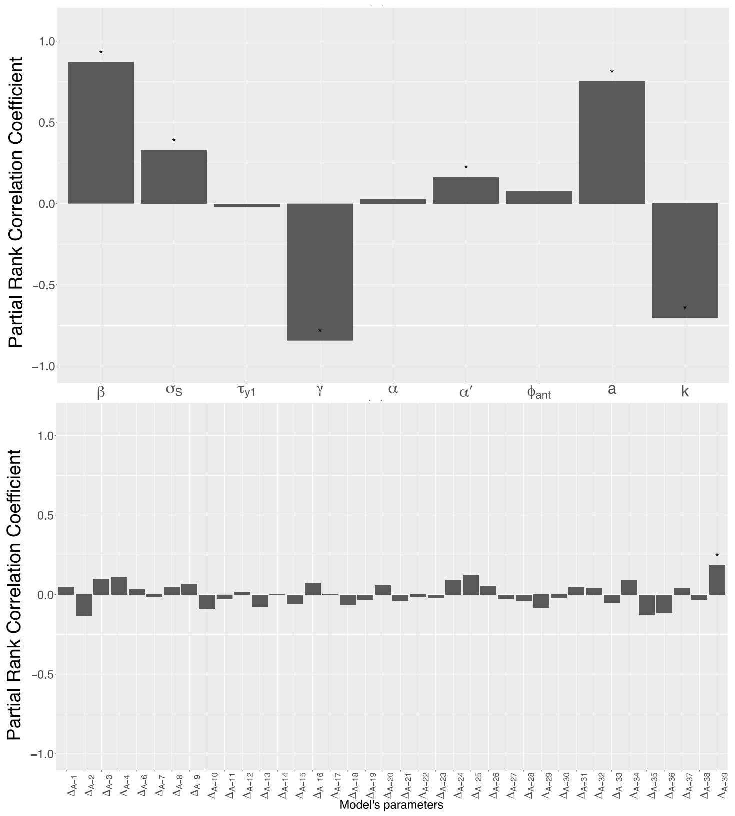


**Emergence case**

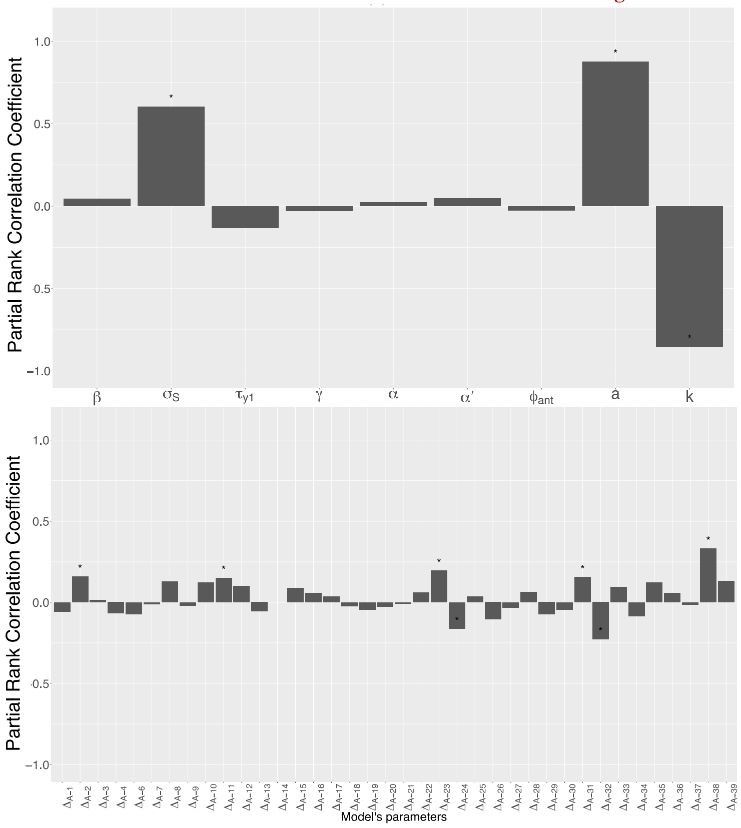

#### (B) Criteria 2 (number of antibiotic-resistant infections) evaluated for country B

##### Endemic case

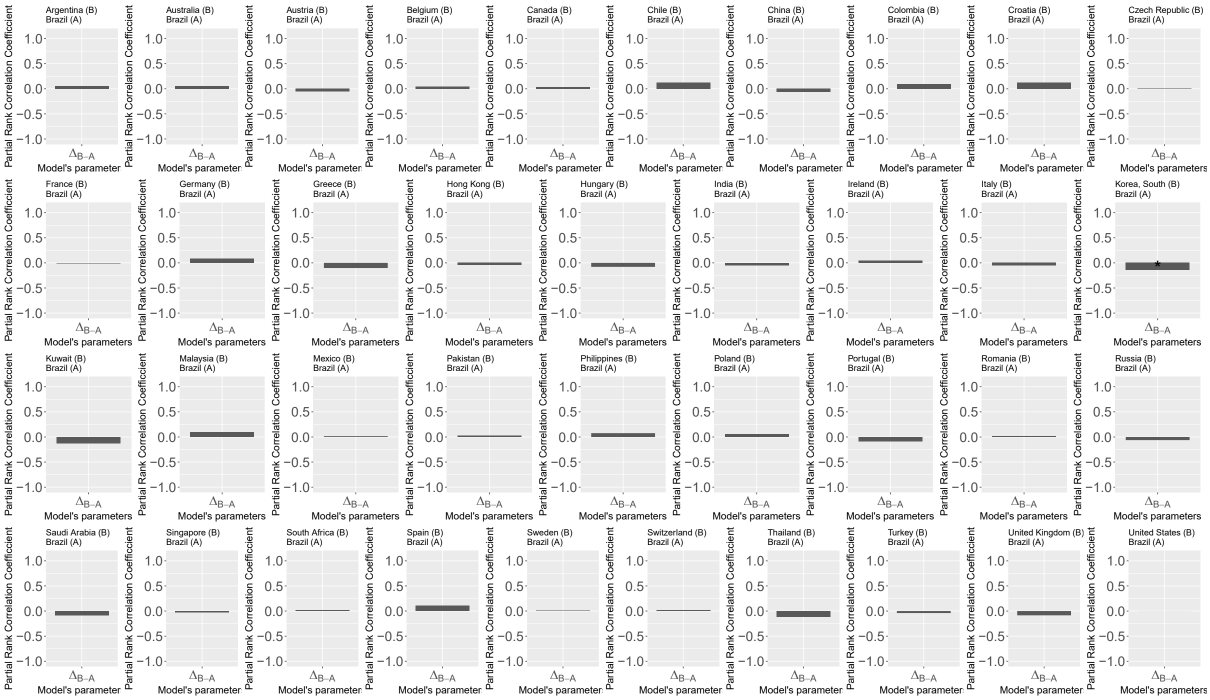


##### Emergence case

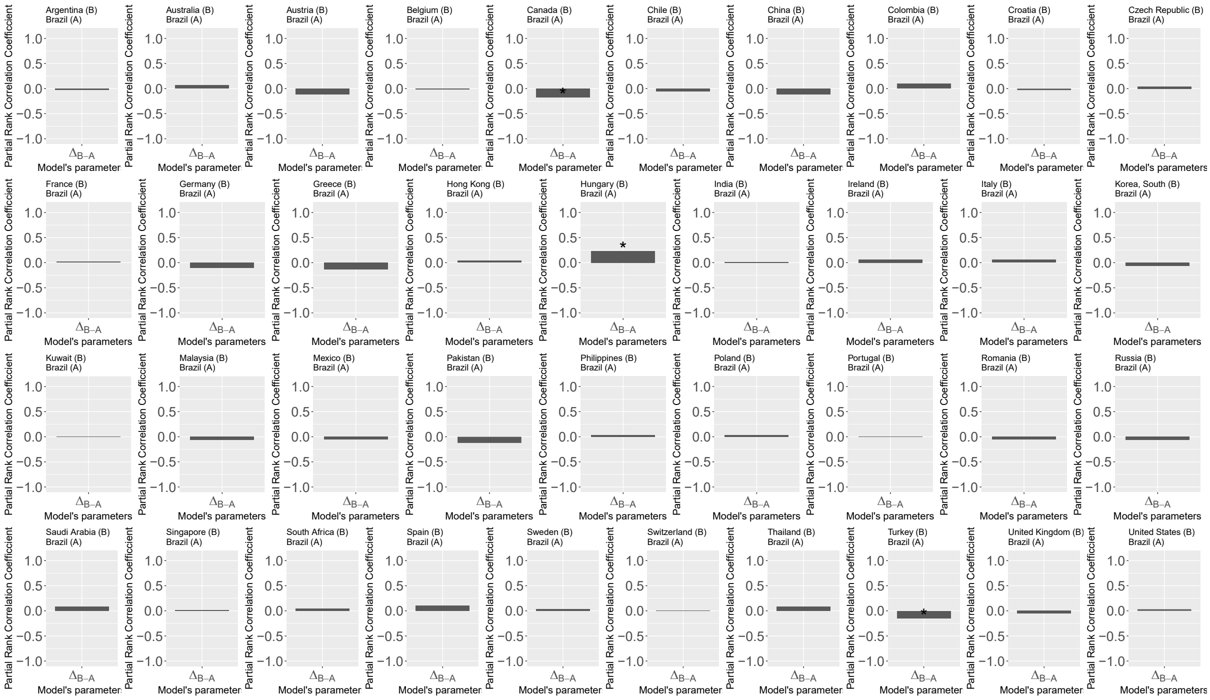


***Figure S5. Sensitivity analysis results: impacts of input parameter uncertainty on the number of antibiotic-resistant infections.***

**Partial rank correlation coefficient (PRCC), for the number of antibiotic-resistant infections (criteria 2) for one country and one year – country A: Brazil. (A):** PRCC between model parameters associated to country A (within-country parameters and A-to-B travel parameters) and criteria 2 evaluated for country A. **(B):** PRCC between B-to-A travel parameters and criteria 2 evaluated for country B after introduction in country A. * indicates a significant association (5% threshold).

#### 4. Procedure for model calibration and results for model comparison.

**Procedure**

For each model evaluated, parameter posterior distributions were obtained after 100,000 iterations, using a burn-in period of 20,000 iterations and subsampling every 200 iterations. An adaptive Metropolis procedure was used. The best model was selected based on minimization of the deviance information criterion (DIC), defined as $DIC = -4 \times mean(LL)+mode(LL)$, with LL the log-likelihood.

**Results**

First, we observe that the inclusion of travel between models 1 and 2 did not significantly reduce the value of the DIC (Figure 3, A). In model 3, the estimated value of parameter $\phi_{ant}$ was close to 0 and did not allow us to quantify a significant impact of antibiotic exposure on the acquisition of ESBL-producing *E. coli*. Model 4, through the country-dependent estimation of the parameter $a_i$, is associated with a slightly reduced DIC whereas model 5, through the country-dependent estimation of the parameter $\beta_i$, is associated with a considerably reduced DIC. The selected model is model 6, corresponding to the lowest DIC value (DIC = 4991).

#### 5. Country-independent model calibration.

To address the identifiability issue raised by the high number of estimated parameters, we performed a country-by-country independent estimation of only four parameters – without accounting for the meta-population framework and international mobility, and running ODEs for a single country at a time. We found a similar order of the median posterior distributions' values when estimated either independently and simultaneously (Figure 3 vs supplementary Figure 6). It suggests that between-country comparisons of estimated parameters can be valid even if absolute estimated parameter values can be dependent on the calibration process.

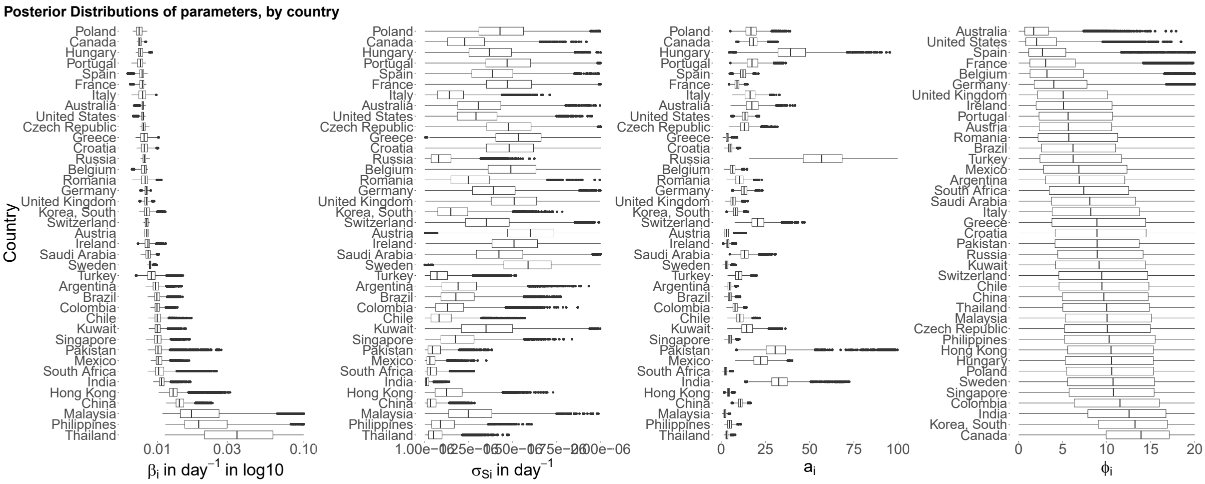


***Figure S6. Parameter estimates for the selected model in a country independent model calibration.***

**Posterior distributions when the four parameters ($\beta_i, a_i, \sigma_{S\,i}, \phi_{ant}$) are estimated independently for each country.** Countries are ordered using median values for $\beta_i$ for the 3 first plots. The order of the countries for median values of $\beta_i$ is mostly the same compared to the order when simultaneous estimations of $\beta_i$ is performed, especially for high median $\beta_i$ values. Only absolute values change. High $a_i$ values estimated independently are also associated with the same countries (Hungary, Russia, Pakistan and India) compared to simultaneous estimations.

#### 6. MCMC results for selected model (model 6).

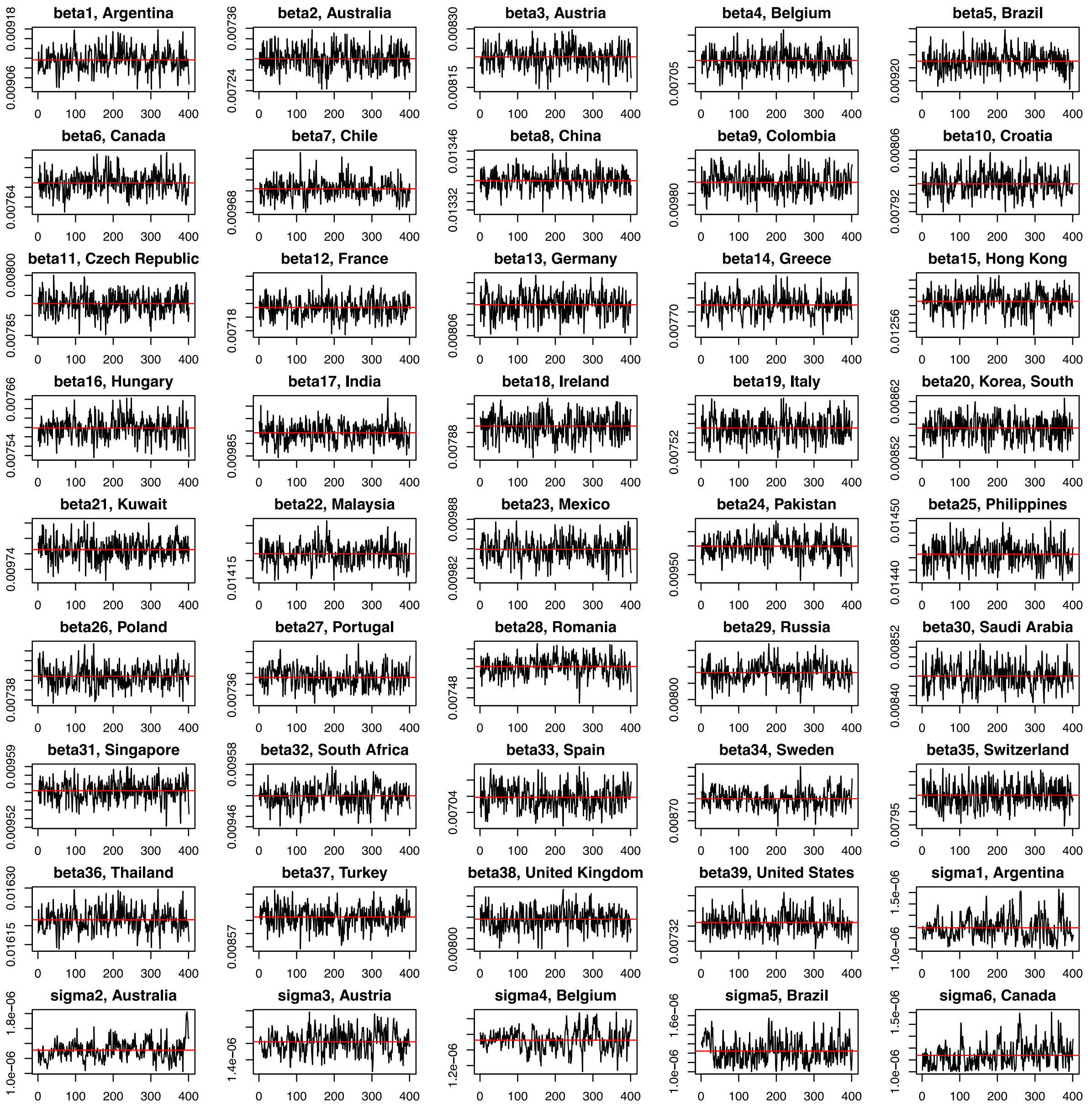

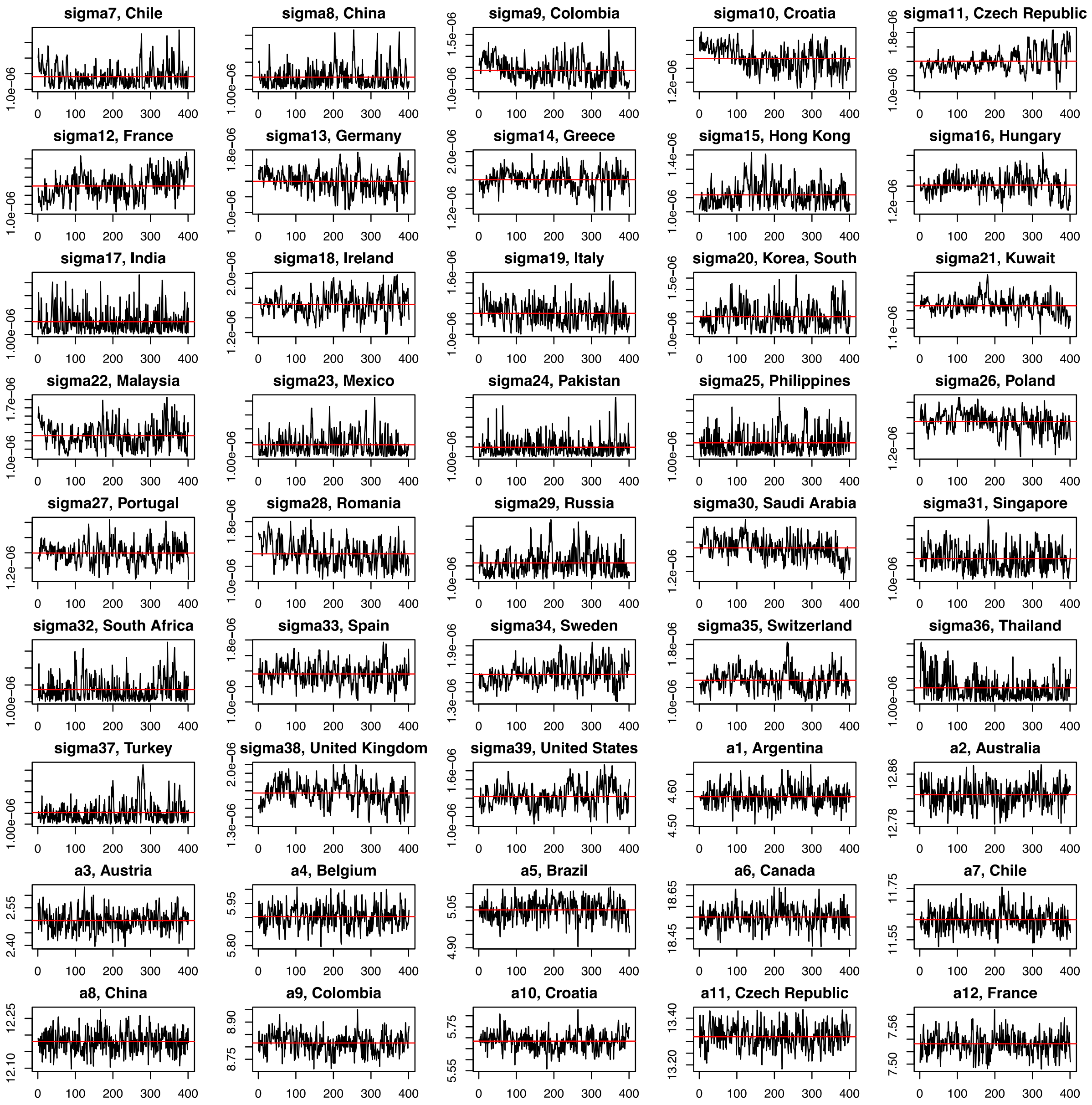
sigma7, Chile
sigma8, China
sigma9, Colombia
sigma10, Croatia
sigma11, Czech Republic
sigma12, France
sigma13, Germany
sigma14, Greece
sigma15, Hong Kong
sigma16, Hungary
sigma17, India
sigma18, Ireland
sigma19, Italy
sigma20, Korea, South
sigma21, Kuwait
sigma22, Malaysia
sigma23, Mexico
sigma24, Pakistan
sigma25, Philippines
sigma26, Poland
sigma27, Portugal
sigma28, Romania
sigma29, Russia
sigma30, Saudi Arabia
sigma31, Singapore
sigma32, South Africa
sigma33, Spain
sigma34, Sweden
sigma35, Switzerland
sigma36, Thailand
sigma37, Turkey
sigma38, United Kingdom
sigma39, United States
a1, Argentina
a2, Australia
a3, Austria
a4, Belgium
a5, Brazil
a6, Canada
a7, Chile
a8, China
a9, Colombia
a10, Croatia
a11, Czech Republic
a12, France

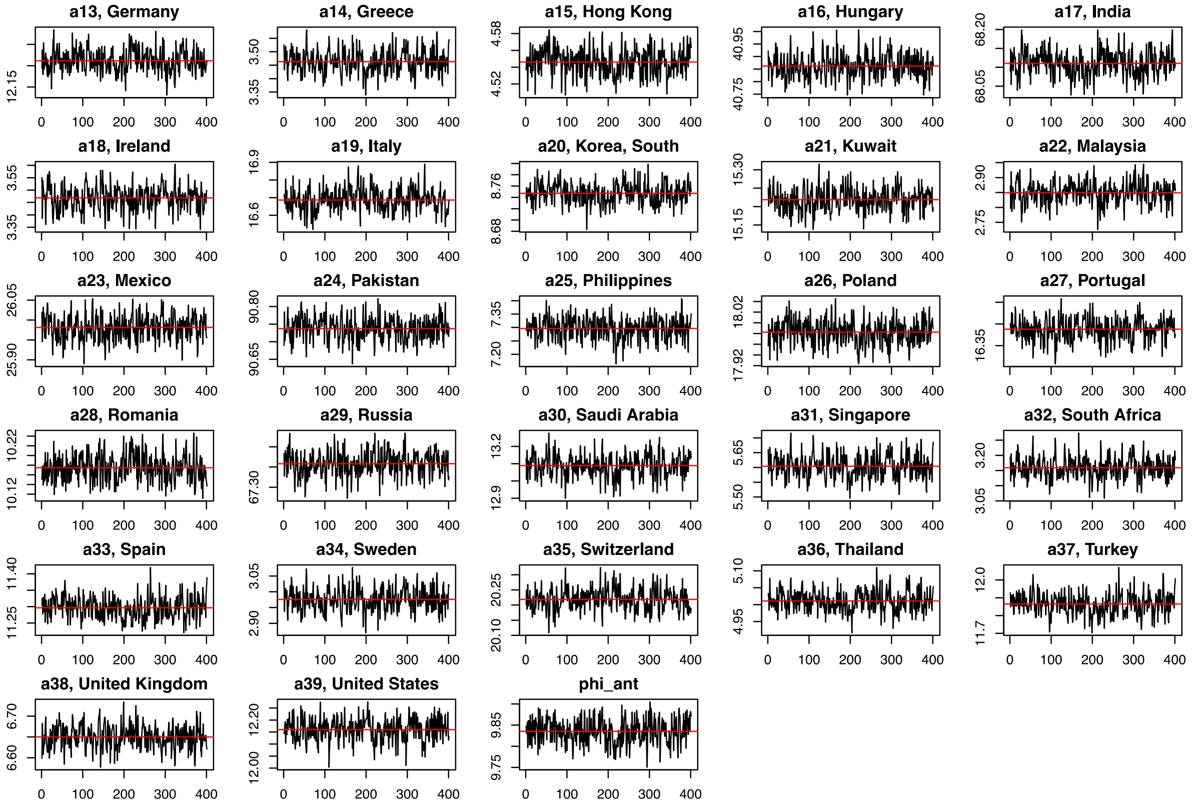


***Figure S7. Traces of the chains obtained for selected model.***

**Traces of accepted parameters values in Metropolis-Hasting for selected model 6.** Traces obtained after 100000 iterations, a 20000 burn-in period and a 200-iterations sampling. Red line indicates median value. $\beta_i, \sigma_{S\,i}, a_i$ (with i from 1 to 39) are the country-dependent parameters and $\phi_{ant}$ is the country independent parameter.

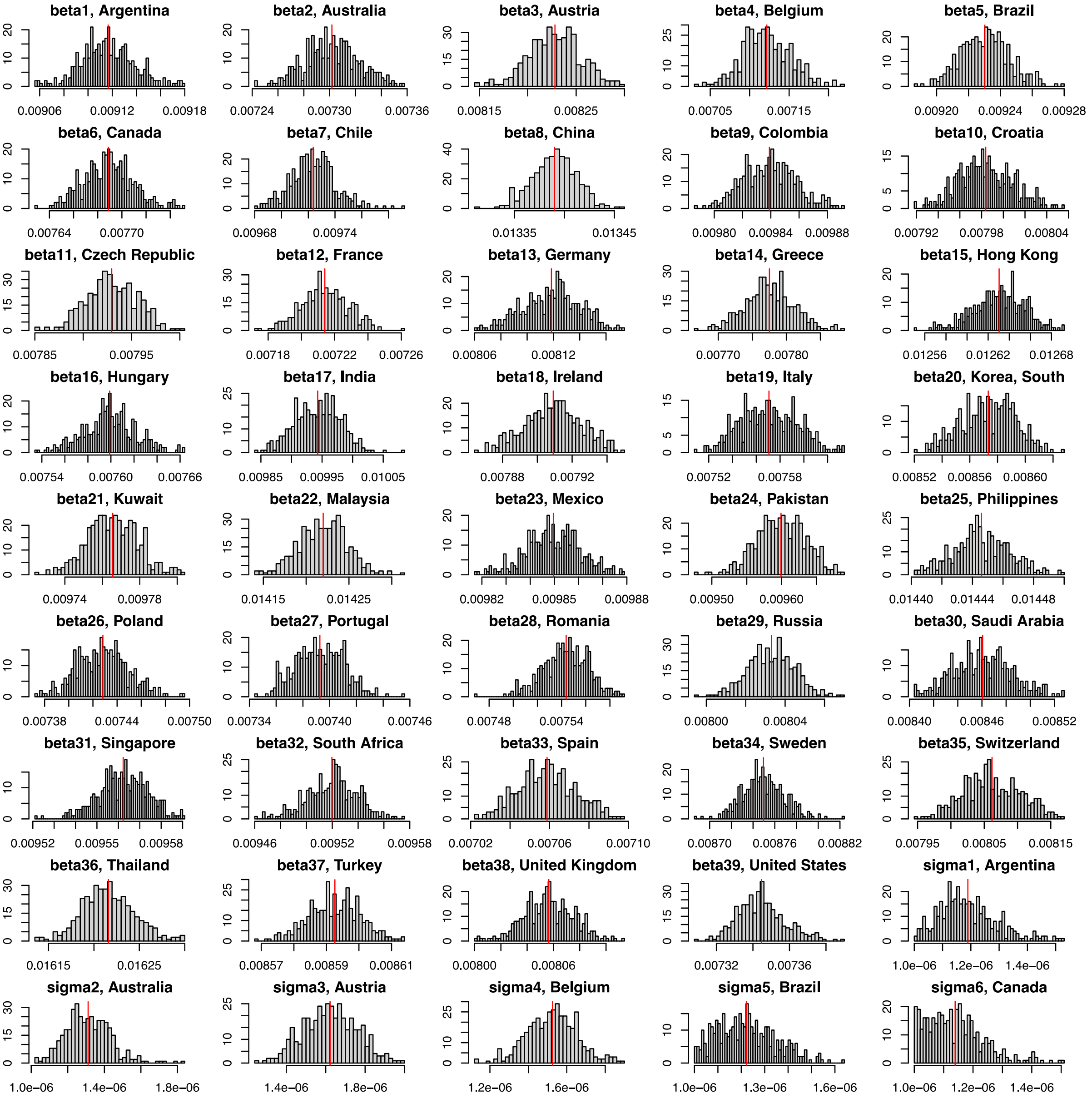
beta1, Argentina
0.00906 0.00912 0.00918
beta2, Australia
0.00724 0.00730 0.00736
beta3, Austria
0.00815 0.00825
beta4, Belgium
0.00705 0.00715
beta5, Brazil
0.00920 0.00924 0.00928
beta6, Canada
0.00764 0.00770
beta7, Chile
0.00968 0.00974
beta8, China
0.01335 0.01345
beta9, Colombia
0.00980 0.00984 0.00988
beta10, Croatia
0.00792 0.00798 0.00804
beta11, Czech Republic
0.00785 0.00795
beta12, France
0.00718 0.00722 0.00726
beta13, Germany
0.00806 0.00812
beta14, Greece
0.00770 0.00780
beta15, Hong Kong
0.01256 0.01262 0.01268
beta16, Hungary
0.00754 0.00760 0.00766
beta17, India
0.00985 0.00995 0.01005
beta18, Ireland
0.00788 0.00792
beta19, Italy
0.00752 0.00758
beta20, Korea, South
0.00852 0.00856 0.00860
beta21, Kuwait
0.00974 0.00978
beta22, Malaysia
0.01415 0.01425
beta23, Mexico
0.00982 0.00985 0.00988
beta24, Pakistan
0.00950 0.00960
beta25, Philippines
0.01440 0.01444 0.01448
beta26, Poland
0.00738 0.00744 0.00750
beta27, Portugal
0.00734 0.00740 0.00746
beta28, Romania
0.00748 0.00754
beta29, Russia
0.00800 0.00804
beta30, Saudi Arabia
0.00840 0.00846 0.00852
beta31, Singapore
0.00952 0.00955 0.00958
beta32, South Africa
0.00946 0.00952 0.00958
beta33, Spain
0.00702 0.00706 0.00710
beta34, Sweden
0.00870 0.00876 0.00882
beta35, Switzerland
0.00795 0.00805 0.00815
beta36, Thailand
0.01615 0.01625
beta37, Turkey
0.00857 0.00859 0.00861
beta38, United Kingdom
0.00800 0.00806
beta39, United States
0.00732 0.00736
sigma1, Argentina
1.0e-06 1.2e-06 1.4e-06
sigma2, Australia
1.0e-06 1.4e-06 1.8e-06
sigma3, Austria
1.4e-06 1.8e-06
sigma4, Belgium
1.2e-06 1.6e-06
sigma5, Brazil
1.0e-06 1.3e-06 1.6e-06
sigma6, Canada
1.0e-06 1.2e-06 1.4e-06

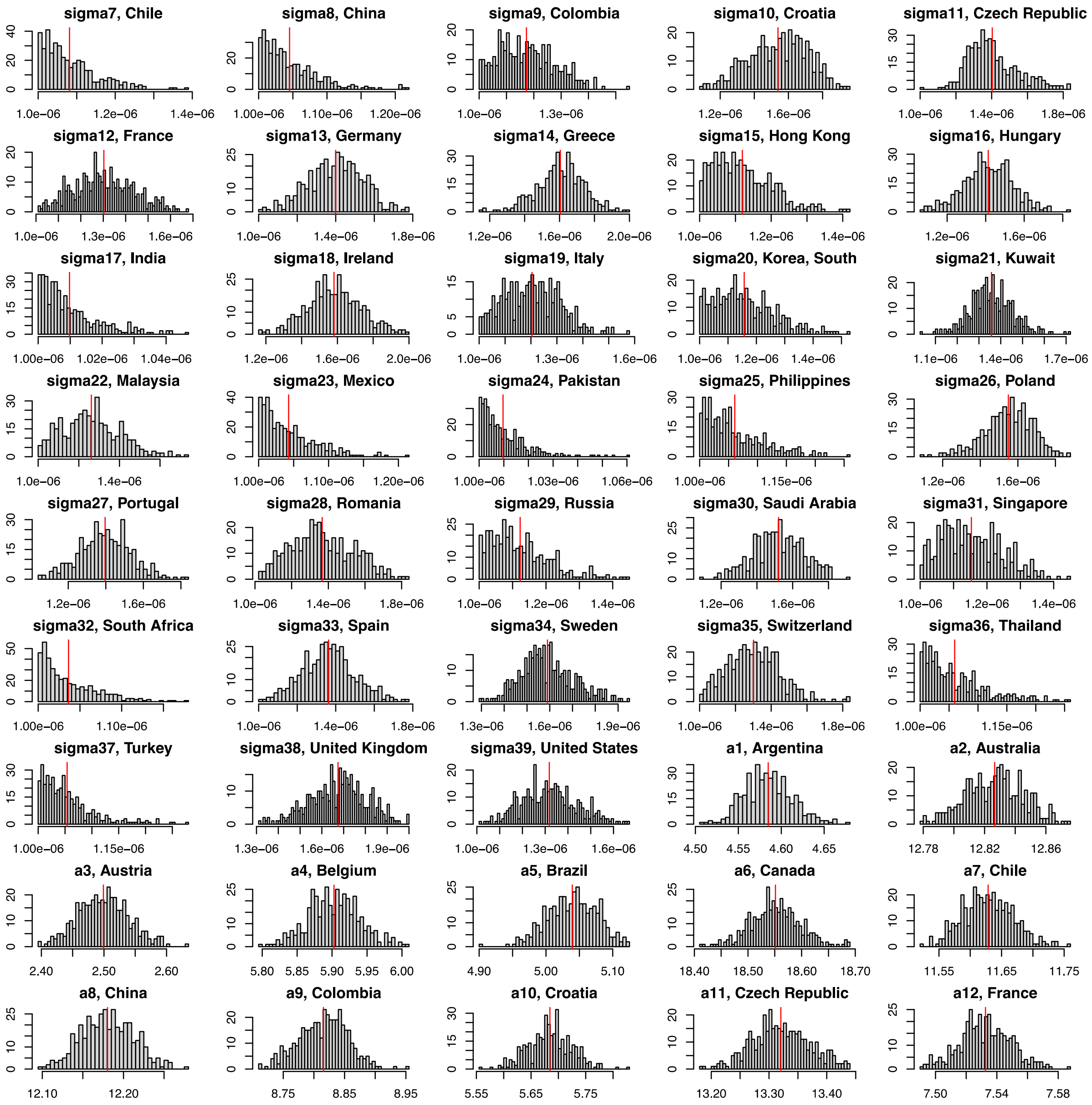
sigma7, Chile
1.0e−06 1.2e−06 1.4e−06
sigma8, China
1.00e−06 1.10e−06 1.20e−06
sigma9, Colombia
1.0e−06 1.3e−06
sigma10, Croatia
1.2e−06 1.6e−06
sigma11, Czech Republic
1.0e−06 1.4e−06 1.8e−06
sigma12, France
1.0e−06 1.3e−06 1.6e−06
sigma13, Germany
1.0e−06 1.4e−06 1.8e−06
sigma14, Greece
1.2e−06 1.6e−06 2.0e−06
sigma15, Hong Kong
1.0e−06 1.2e−06 1.4e−06
sigma16, Hungary
1.2e−06 1.6e−06
sigma17, India
1.00e−06 1.02e−06 1.04e−06
sigma18, Ireland
1.2e−06 1.6e−06 2.0e−06
sigma19, Italy
1.0e−06 1.3e−06 1.6e−06
sigma20, Korea, South
1.0e−06 1.2e−06 1.4e−06
sigma21, Kuwait
1.1e−06 1.4e−06 1.7e−06
sigma22, Malaysia
1.0e−06 1.4e−06
sigma23, Mexico
1.00e−06 1.10e−06 1.20e−06
sigma24, Pakistan
1.00e−06 1.03e−06 1.06e−06
sigma25, Philippines
1.00e−06 1.15e−06
sigma26, Poland
1.2e−06 1.6e−06
sigma27, Portugal
1.2e−06 1.6e−06
sigma28, Romania
1.0e−06 1.4e−06 1.8e−06
sigma29, Russia
1.0e−06 1.2e−06 1.4e−06
sigma30, Saudi Arabia
1.2e−06 1.6e−06
sigma31, Singapore
1.0e−06 1.2e−06 1.4e−06
sigma32, South Africa
1.00e−06 1.10e−06
sigma33, Spain
1.0e−06 1.4e−06 1.8e−06
sigma34, Sweden
1.3e−06 1.6e−06 1.9e−06
sigma35, Switzerland
1.0e−06 1.4e−06 1.8e−06
sigma36, Thailand
1.00e−06 1.15e−06
sigma37, Turkey
1.00e−06 1.15e−06
sigma38, United Kingdom
1.3e−06 1.6e−06 1.9e−06
sigma39, United States
1.0e−06 1.3e−06 1.6e−06
a1, Argentina
4.50 4.55 4.60 4.65
a2, Australia
12.78 12.82 12.86
a3, Austria
2.40 2.50 2.60
a4, Belgium
5.80 5.85 5.90 5.95 6.00
a5, Brazil
4.90 5.00 5.10
a6, Canada
18.40 18.50 18.60 18.70
a7, Chile
11.55 11.65 11.75
a8, China
12.10 12.20
a9, Colombia
8.75 8.85 8.95
a10, Croatia
5.55 5.65 5.75
a11, Czech Republic
13.20 13.30 13.40
a12, France
7.50 7.54 7.58

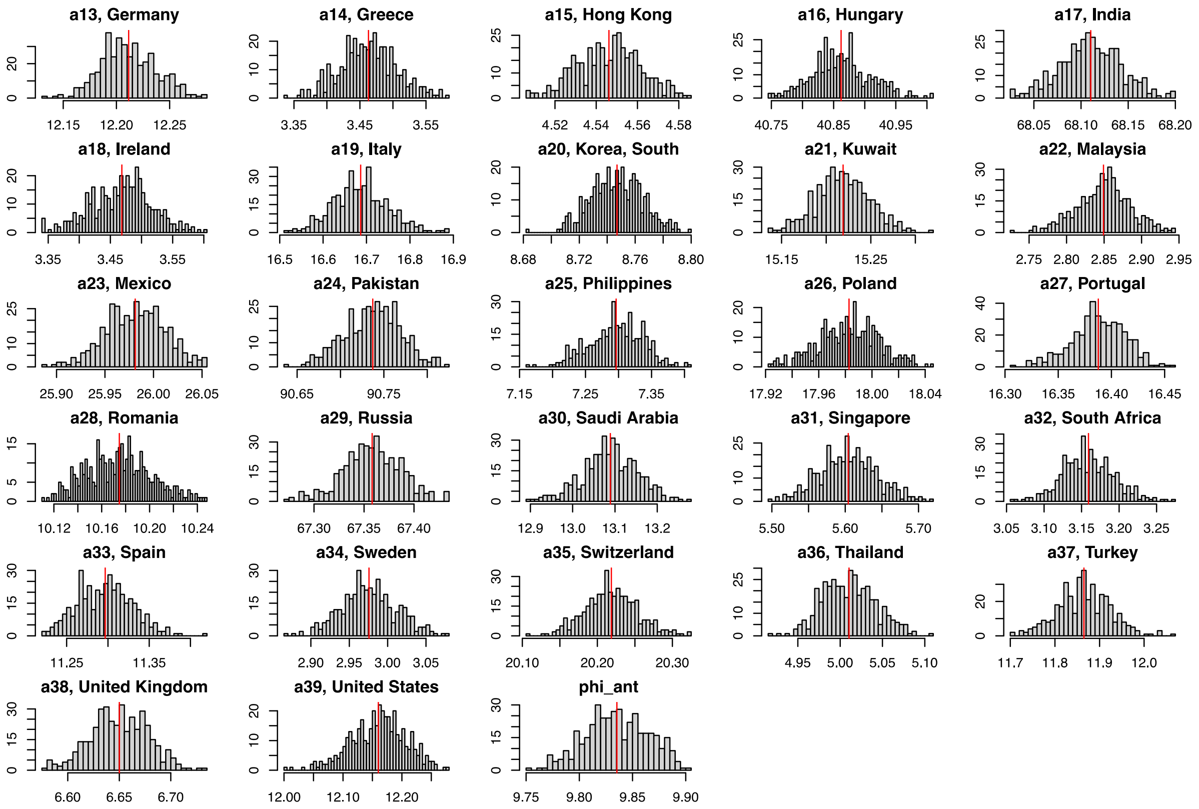


***Figure S8. Posterior distributions for selected model.***

**Posterior distributions of accepted values in Metropolis-Hasting for selected model 6.** Distributions obtained after 100000 iterations, a 20000 burn-in period and a 200-iterations sampling. Red line indicates the median value. $\beta_i, \sigma_{S\,i}, a_i$ (with i from 1 to 39) are the country-dependent parameters and $\phi_{ant}$ is the country independent parameter.

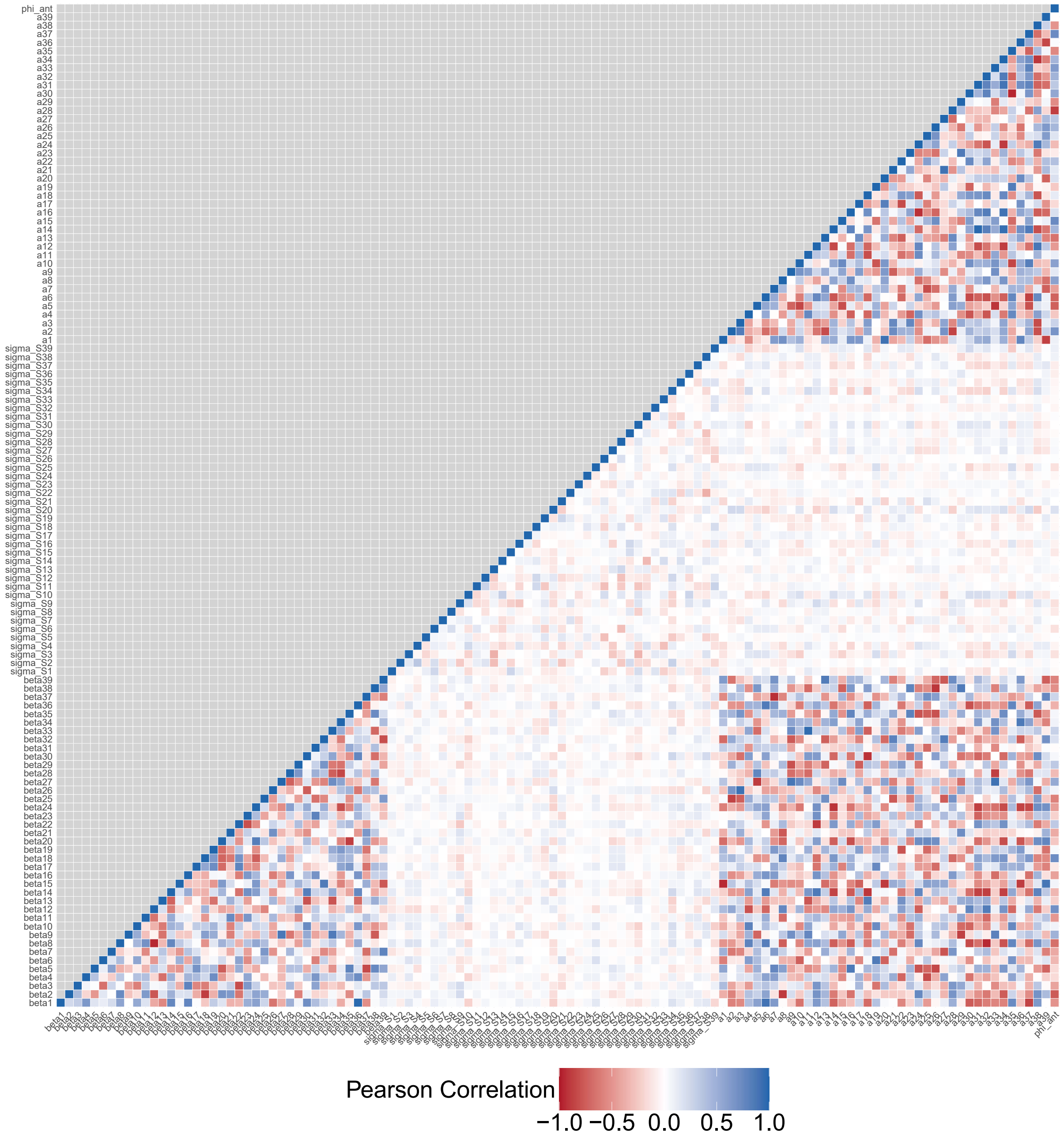


***Figure S9. Parameter correlation in the MCMC chains with the simultaneous fit approach.***

Correlation coefficients between median MCMC accepted values for simultaneously estimated parameters.

A.

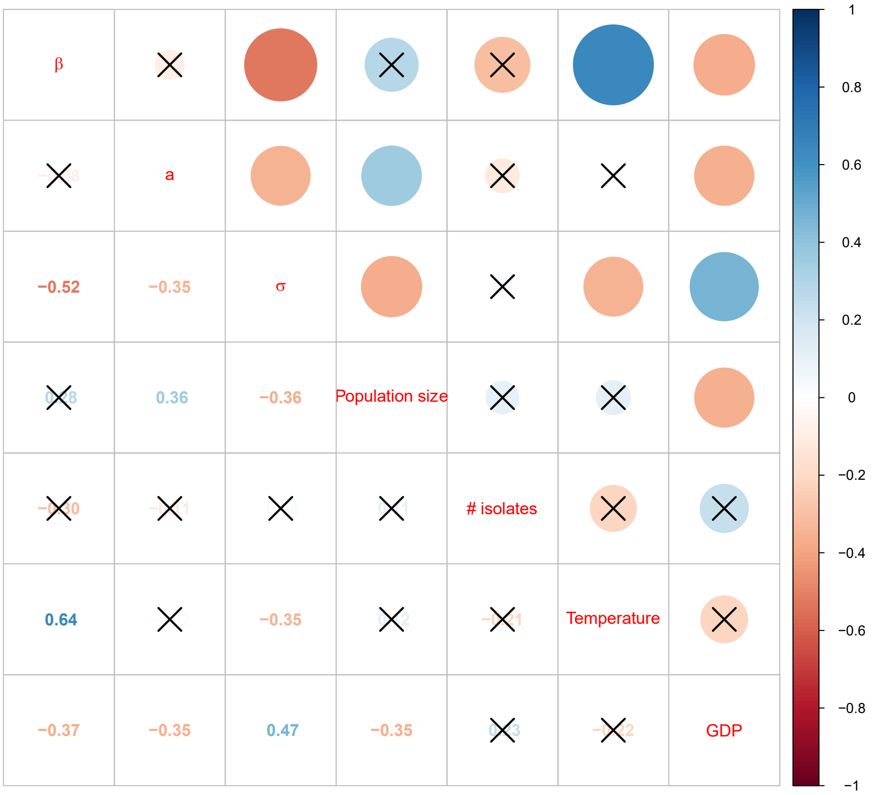

β
a
σ
Population size
### isolates
Temperature
GDP
−0.52
−0.35
0.36
−0.36
0.64
−0.35
−0.37
−0.35
0.47
−0.35
1
0.8
0.6
0.4
0.2
0
−0.2
−0.4
−0.6
−0.8
−1


B.

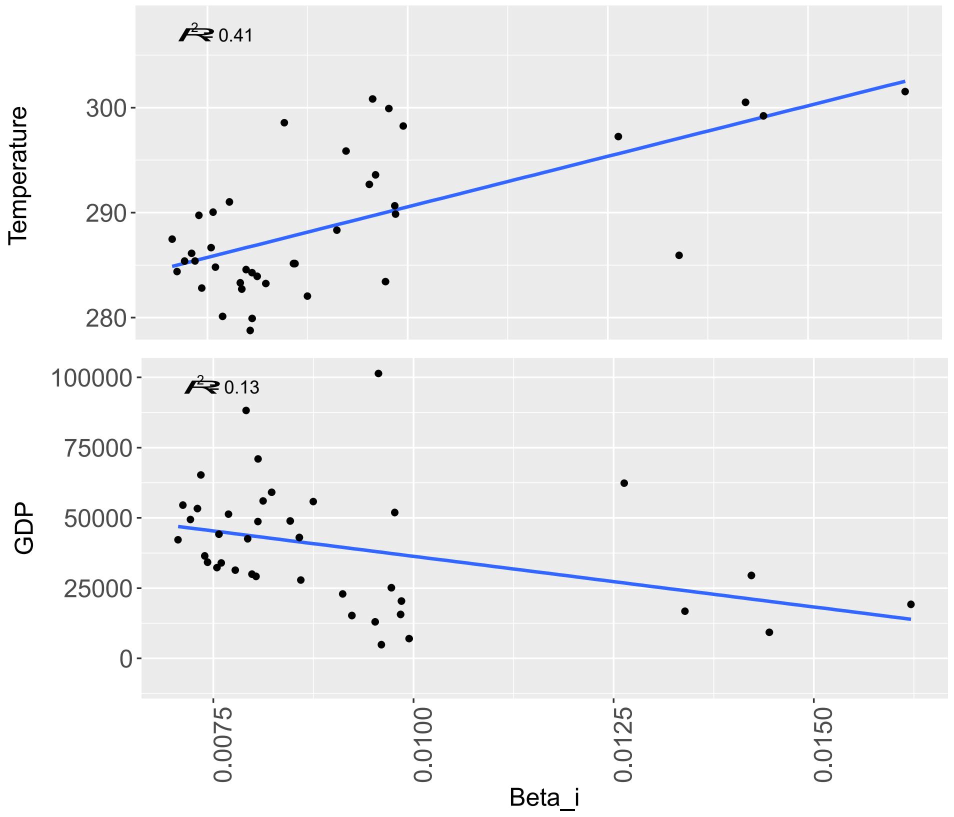

$R^2$ 0.41
Temperature
300
290
280
$R^2$ 0.13
GDP
100000
75000
50000
25000
0
0.0075
0.0100
0.0125
0.0150
Beta_i

***Figure S10. Correlations between median country-dependent parameter values obtained from posterior distributions (model 6) and other country-dependent factors.***

**(A):** Pearson correlation coefficients (r). Cross indicates a non-significant association. # isolates: number of isolates reported by country to the ATLAS reporting system. **(B):** Linear associations between $\beta_i$ and temperature (in Kelvin) or gross domestic product (GDP) with Pearson correlation coefficients r = 0,64 [0,41-0,79] (p-value < $1x10^{-5}$) for temperature and r = -0,36 [-0,61-0,06] (p-value = 0,02) for GDP.
